# Clustering of patient comorbidities within electronic medical records enables high-precision COVID-19 mortality prediction

**DOI:** 10.1101/2021.03.29.21254579

**Authors:** Erwann Le Lannou, Benjamin Post, Shlomi Haar, Stephen J. Brett, Balasundaram Kadirvelu, A. Aldo Faisal

## Abstract

We present an explainable AI framework to predict mortality after a positive COVID-19 diagnosis based solely on data routinely collected in electronic healthcare records (EHRs) obtained prior to diagnosis. We grounded our analysis on the ½ Million people UK Biobank and linked NHS COVID-19 records. We developed a method to capture the complexities and large variety of clinical codes present in EHRs, and we show that these have a larger impact on risk than all other patient data but age. We use a form of clustering for natural language processing of the clinical codes, specifically, topic modelling by Latent Dirichlet Allocation (LDA), to generate a succinct digital fingerprint of a patient’s full secondary care clinical history, i.e. their comorbidities and past interventions. These digital comorbidity fingerprints offer immediately interpretable clinical descriptions that are meaningful, e.g. grouping cardiovascular disorders with common risk factors but also novel groupings that are not obvious. The comorbidity fingerprints differ in both their breadth and depth from existing observational disease associations in the COVID-19 literature. Taking this data-driven approach allows us to avoid human-induction bias and confirmation bias during selection of what are important potential predictors of COVID-19 mortality. Together with age, these digital fingerprints are the single most important factor in our predictor. This holds the potential for improving individual risk profiling for clinical decisions and the identification of groups for public health interventions such as vaccine programmes. Combining our digital precondition fingerprints with demographic characteristics allow us to match or exceed the performance of existing state-of-the-art COVID-19 mortality predictors (EHCF) which have been developed through expert consensus. Our precondition fingerprinting and entire mortality prediction analytics pipeline are designed so as to be rapidly redeployable, e.g. for COVID-19 variants or other pre-existing diseases.

## Introduction

The outbreak of the novel, severe and acute respiratory syndrome, Coronavirus 2 (SARS-CoV-2), and its associated disease COVID-19, in Wuhan China, has presented an important and urgent threat to global health since December 2019. Declared a pandemic by the World Health Organisation (WHO) in early March^1^, this disease has, as of 8 March 2021, caused 2,582,528 deaths from 116,166,652 cumulative worldwide cases^2^. On the same day in the United Kingdom (UK) SARS-CoV-2 is reported as responsible for 124,419 deaths^2^. The COVID-19 outbreak has led to an increase in hospital admissions and has considerably increased the demand for both general hospital and critical care beds^3, 4^. This increases the need for an objective, data-driven understanding of risk factors and accurate prediction of adverse outcomes based on pre-existing information. Such understanding may facilitate better individual clinical decisions, but potentially also increase the resilience of public health policy.

Emerging evidence throughout the pandemic has identified several risk factors associated with COVID-19 clinical severity^5^ and fatality^6, 7^. Most commonly reported risk factors are old age^8–10^, male gender^11, 12^ and pre-existing underlying medical conditions^6^. In particular, the Chinese Centre for Disease Control and Prevention identified cardiovascular disease, hypertension, diabetes, respiratory disease, and numerous malignancies to be risk factors for COVID-19 fatality^12^. In the United States, the most commonly initially reported comorbidities were hypertension, obesity and diabetes^9^. Finally, a UK-wide study aiming to characterise the clinical features of patients with severe COVID-19 highlighted chronic cardiac disease, diabetes, and chronic pulmonary disease to be associated with a higher risk of clinical severity^13–15^. Better knowledge of an individual’s risk can help better prioritise risk groups in the global vaccination effort as well as tailor our efforts for both prevention (e.g. targeted isolation or social distancing) and for treatment of confirmed cases such as immediate hospitalisation for people at greater risk. Many attempts have been made to provide risk prediction models for COVID-19. Already in April 2020, Wynants et al. reviewed 145 prediction models^16^. Of the 145 described models, 23 models were aimed at estimating the risk of mortality. The review further identified that most studies had been performed on inpatients and required lab-testing (lymphocyte count, C-reactive protein, creatinine) and so were not aimed at community-based identification. In addition, the review concluded that the currently proposed models were ‘poorly reported, at high risk of bias, and their reported performance is probably optimistic’, with sample sizes rarely over 1000^16^. In order to define who is at high risk, governmental organisations have put forward defining criteria that aim to classify low- and high-risk groups of the population^17, 18^. However, at present these criteria are developed by ‘expert consensus’ based on available bodies of evidence and so require time and effort and might be biased by the experts’ priors. While the co-morbidities have been observed in affecting COVID-19 mortality^19^ they have not been systematically analysed for building a principled integrated COVID-19 mortality risk predictor. Preconditions, especially if they appear grouped, so called multi-morbidities affect a substantial proportion of the world’s population^20^. Estimates of prevalence vary depending on population examined and definitions used, in the UK >50% of people have at least two conditions by age 65, and >50% have 3+ conditions by age 70^21^. Hence, we took a more principled approach to incorporating patient specific information by systematically including comorbidities and their co-occurrence in our data-driven analysis to avoid human-induction bias, while carefully avoiding and signposting the potential for AI bias further below^22^. Our approach is enabled by the existence of large population datasets with linkage to registries such as death records, hospital admission and COVID-19 testing results and therefore represent a novel opportunity for automated clinical risk prediction model development on larger cohorts. Here, we used the UK Biobank^23, 24^, which since the 16 March 2020 has linked over 26,000 COVID-19 laboratory test results electronically to its 500,000 strong cohort of participants. The use of such linked datasets has an established track record for the development and evaluation of clinical risk models, including those for cardiovascular disease, cancer, and mortality^25–28^.

In this paper, we utilise the longitudinal medical records from the UK Biobank to predict COVID-19-related death. We demonstrate that the data already routinely collected and stored in EHRs can be rapidly leveraged to address pressing questions related to the COVID-19 pandemic. Notably, we apply a Topic Modelling approach, Latent Dirichlet Allocation (LDA), on the entirety of the UK Biobank population to construct precise ‘digital comorbidity fingerprints’ (DCFs) from EHR data. Using the early data from the UK COVID-19 epidemic, we explored the utility of our approach by using the DCFs to train a Random Forrest Classifier to predict COVID-19-related death. We demonstrate the face validity of such a two-step approach to model development by comparing the prediction performance of our DCFs, combined with previously documented demographic data, to a model based on expert consensus led feature selection, and one based entirely on age. We further consider the relationship between our data-driven DCFs and clinician-driven features. The approach proposed here is designed to provide a rapid, intuitive and accurate forecasting of COVID-19-related death based on past medical records, which is of special importance for managing hospital resources, vaccination efforts and preventative policies.

## Results

### Characteristics of the study population

A total of 2,499 COVID-19 patients were included in this analysis (Figure 1; cohort description in Table 1). Overall, the median (IQR) age of patients at baseline was 70.4 (59.9 - 76.3, range: 50.4 - 83.7) years, and 1,284 patients (51.4 %) were male. Over the study period, COVID-19-related deaths were recorded in linked death registration data for 349 patients (14.0 % of the study population). The hospitalised patients encompassed 79% of the 349 death events. Figure 1, shows the weekly number of positive test results during the study period, from January 30 to September 9, along with the weekly number of COVID-19 related deaths in the same period. We can observe a peak in early April 2020 (week of the 8 April, 272 people tested positive) followed by a peak in COVID-19 related deaths a week later (week of the 15 April 2020, 61 COVID-19 related deaths). The median age of the patients who died of a COVID-19 related death was 76.0 years (76.1 years for men and 75.4 years for women). The median follow-up time for all patients (time between diagnosis and either death or end of the study) was 113 days (10 days for COVID-19-related deaths and 123 days for patients still alive at the end of the study period). Figure 1 shows the overall probability of survival 28 days after diagnosis was over 0.88, dropping to 0.82; whereas for participants over 80 years old, the survival probability was 0.69 after 28 days falling to 0.64.

**Figure 1.**
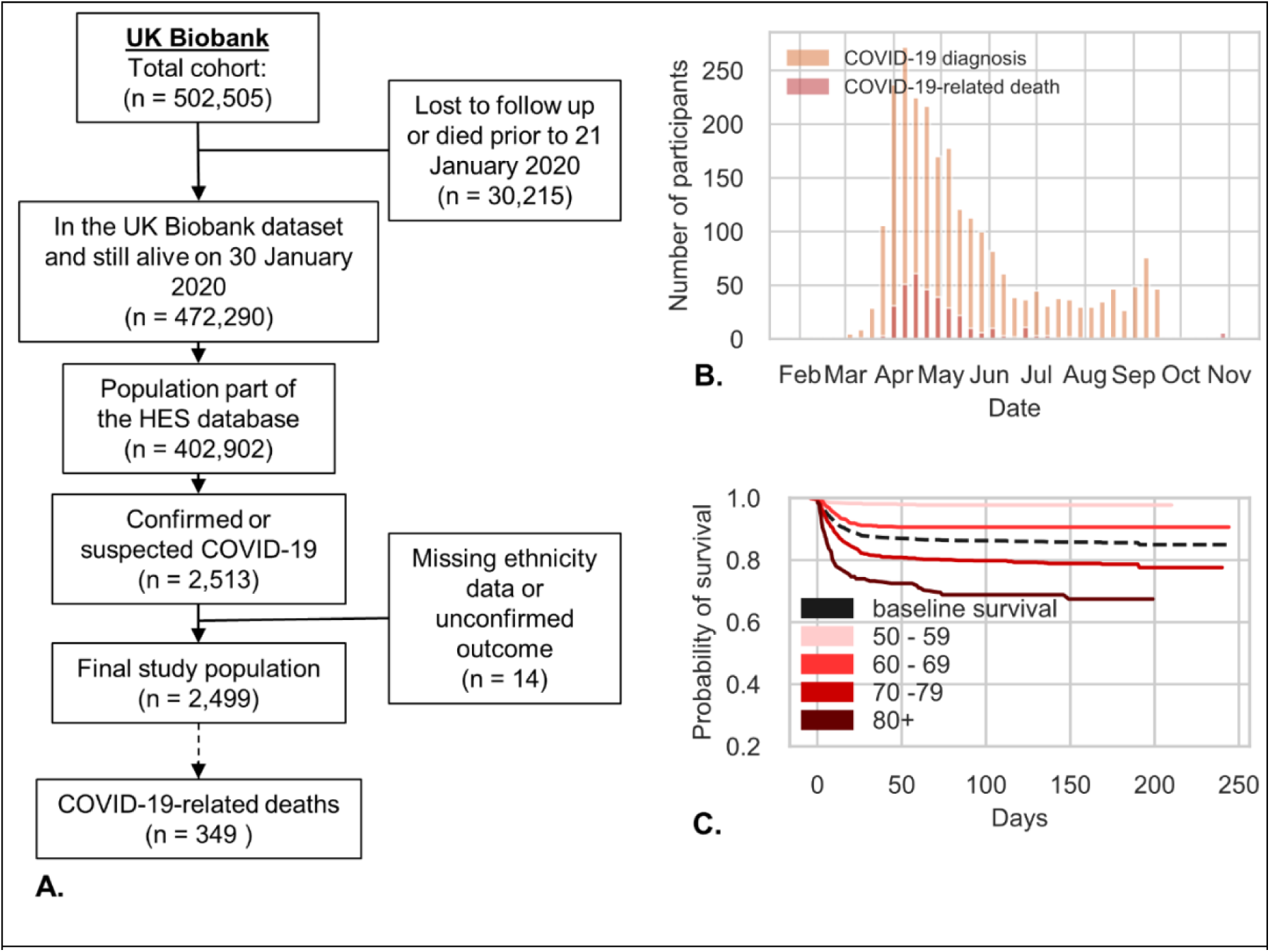
Cohort description. (A) Flowchart diagram of the study cohort. (B) Distribution of COVID-19 diagnosis and death. (C) Kaplan-Meier plots for COVID-19-related death per age group

**Table 1.** Descriptive characteristics of the UK Biobank cohort by COVID-19 fatality and Odds ratios and 95 % confidence intervals for COVID-19-related death

|  | **COVID-19-**<br>**related death**<br>**(n = 349)** | **Rest of the**<br>**Cohort (n =**<br>**2150)** | **OR (95% CI)** | **Fully adjusted OR**<br>**(95% CI)** |
| --- | --- | --- | --- | --- |
| Age 50-59 | 14 (4.01%) | 619<br>(28.79%) | 1 | 1 |
| Age 60-69 | 55 (15.76%) | 537<br>(24.98%) | 4.53 (2.49 - 8.23) | 3.56 (1.93 - 6.54) |
| Age 70-79 | 234 (67.05%) | 894<br>(41.58%) | 11.57 (6.68 - 20.04) | 8.00 (4.49 - 14.25) |
| Age 80+ | 46 (13.18%) | 100 (4.65%) | 20.34 (10.78 - 38.36) | 14.03 (7.18 - 27.44) |
| Sex | 221 (63.32%) | 1063<br>(49.44%) | 1.77 (1.40 - 2.23) | 1.50 (1.16 - 1.94) |
| Deprivation Q1 | 64 (18.34%) | 389<br>(18.09%) | 1 | 1 |
| Deprivation Q2 | 69 (19.77%) | 427<br>(19.86%) | 0.98 (0.68 - 1.42) | 0.96 (0.65 - 1.41) |
| Deprivation Q3 | 80 (22.92%) | 567<br>(26.37%) | 0.86 (0.60 - 1.22) | 0.86 (0.59 - 1.25) |
| Deprivation Q4 | 136 (38.97%) | 767<br>(35.67%) | 1.08 (0.78 - 1.49) | 1.11 (0.78 - 1.57) |
| Ethnicity (non-white) | 35 (10.03%) | 243<br>(11.30%) | 0.87 (0.60 - 1.27) | 1.40 (0.91 - 2.14) |
| Obesity | 67 (19.20%) | 279<br>(12.98%) | 1.59 (1.19 - 2.14) | 1.24 (0.89 - 1.74) |
| Smoking status | 240 (68.77%) | 1326<br>(61.67%) | 1.37 (1.07 - 1.74) | 1.10 (0.84 - 1.44) |
| Organ transplant | 3 (0.86%) | 19 (0.88%) | 0.97 (0.29 - 3.30) | 0.60 (0.17 - 2.17) |
| Cancer | 97 (27.79%) | 432<br>(20.09%) | 1.53 (1.18 - 1.98) | 1.13 (0.86 - 1.48) |
| Severe respiratory disease | 69 (19.77%) | 237<br>(11.02%) | 1.99 (1.48 - 2.67) | 1.03 (0.73 - 1.45) |
| Rare disease or inborn errors of metabolism | 6 (1.72%) | 18 (0.84%) | 2.07 (0.82 - 5.26) | 1.89 (0.71 - 5.04) |
| Congenital heart defects | 4 (1.15%) | 11 (0.51%) | 2.25 (0.71 - 7.12) | 1.74 (0.49 - 6.09) |
| Cardiovascular disease | 206 (59.03%) | 761<br>(35.40%) | 2.63 (2.09 - 3.31) | 1.09 (0.81 - 1.46) |
| Chronic, non-severe respiratory disease | 52 (14.90%) | 302<br>(14.05%) | 1.07 (0.78 - 1.47) | 0.87 (0.61 - 1.24) |
| Chronic kidney disease | 59 (16.91%) | 190 (8.84%) | 2.10 (1.53 - 2.88) | 0.97 (0.68 - 1.40) |
| Cardiac disease | 71 (20.34%) | 211 (9.81%) | 2.35 (1.74 - 3.16) | 1.21 (0.86 - 1.71) |
| Liver disease | 42 (12.03%) | 176 (8.19%) | 1.53 (1.07 - 2.19) | 1.07 (0.73 - 1.59) |
| Chronic neurological conditions | 77 (22.06%) | 215<br>(10.00%) | 2.55 (1.91 - 3.40) | 1.64 (1.20 - 2.24) |
| Diabetes | 100 (28.65%) | 353<br>(16.42%) | 2.04 (1.58 - 2.65) | 1.21 (0.89 - 1.63) |
| Conditions or treatments that predispose to infection | 50 (14.33%) | 157 (7.30%) | 2.12 (1.51 - 2.98) | 1.52 (1.05 - 2.20) |
| Hypertension | 238 (68.19%) | 942<br>(43.81%) | 2.75 (2.16 - 3.50) | 1.34 (1.00 - 1.81) |

In order to better characterise the impact of pre-existing conditions on the study population, we used the categories from our Expert Hand-Crafted Features (EHCF) (see Box 1). The most common comorbidity in the population was hypertension (47.4%) followed by Cardiovascular disease (39.0%) and cancer (21.1%) (Table 1). The univariate and fully adjusted associations (calculated using logistic regression) between patient level characteristics (Table 1) and odds of COVID-19-related death are shown in Table 1 and Figure 2. Increasing age showed the strongest association with an increased likelihood of COVID-19-related death; with participants aged over 80 years being over 15 times (fully adjusted OR: 15.86, 95% CI: 7.85 - 32.04) more likely to die of a COVID-19-related death compared to 50–59-year-olds. Other significant characteristics that showed an increased likelihood of COVID-19-related death are male gender (fully adjusted OR: 1.54, 95% CI: 1.18 - 2.01), and a previous diagnosis of hypertension (fully adjusted OR: 1.54 (1.14 - 2.09)). These findings are consistent with previously reported associations^29–31^.

**Box 1.**
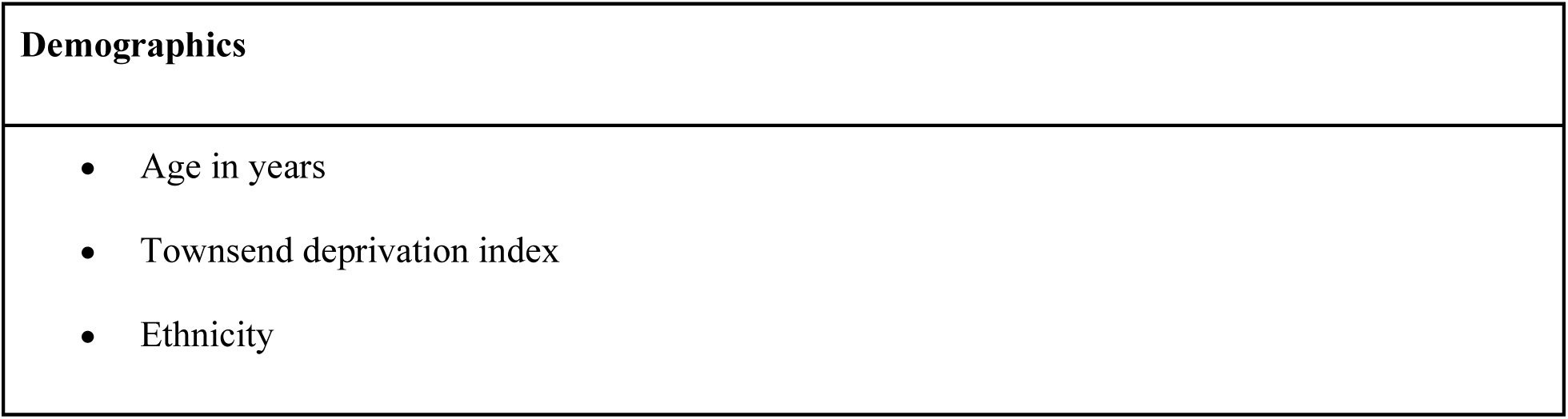
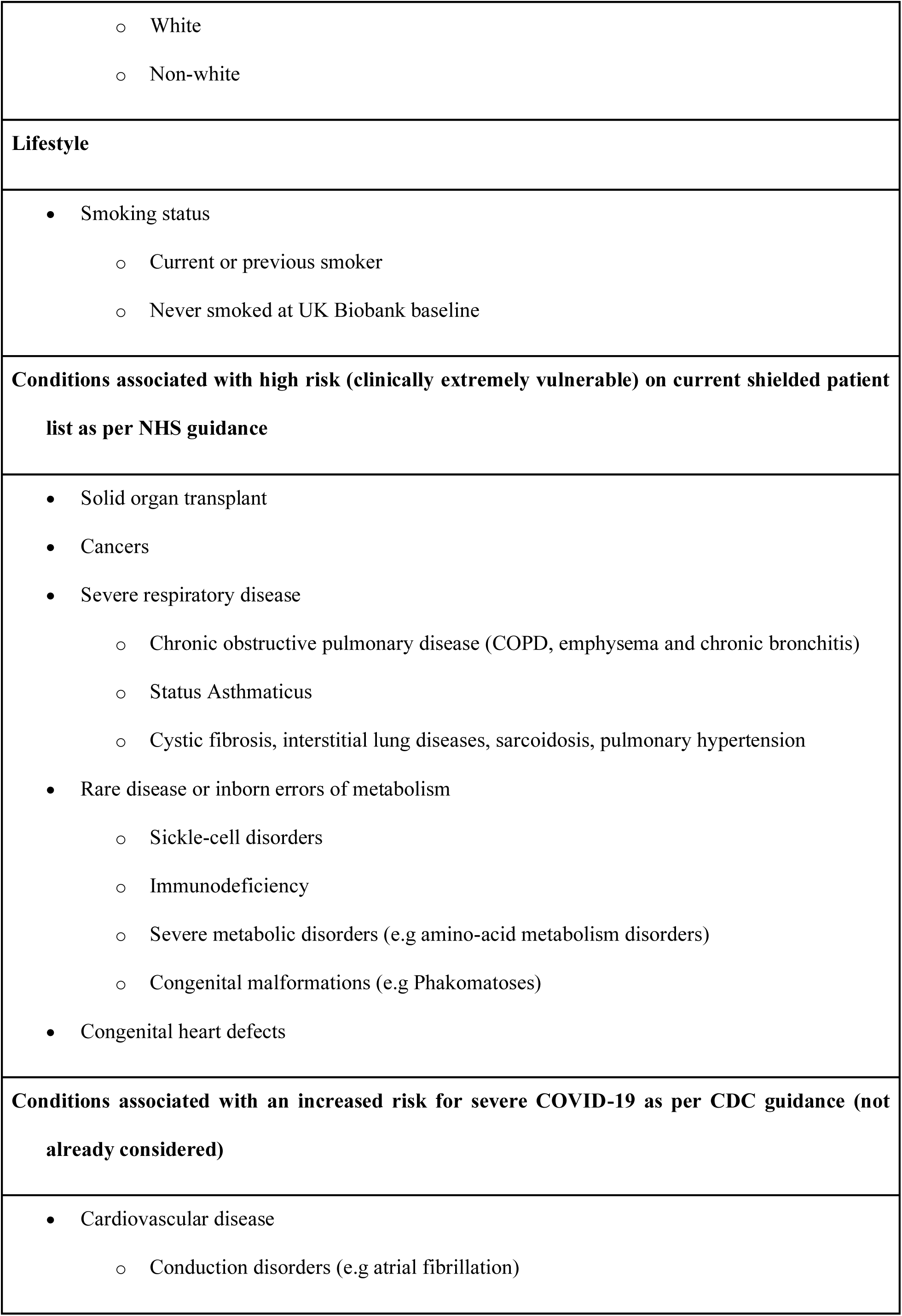
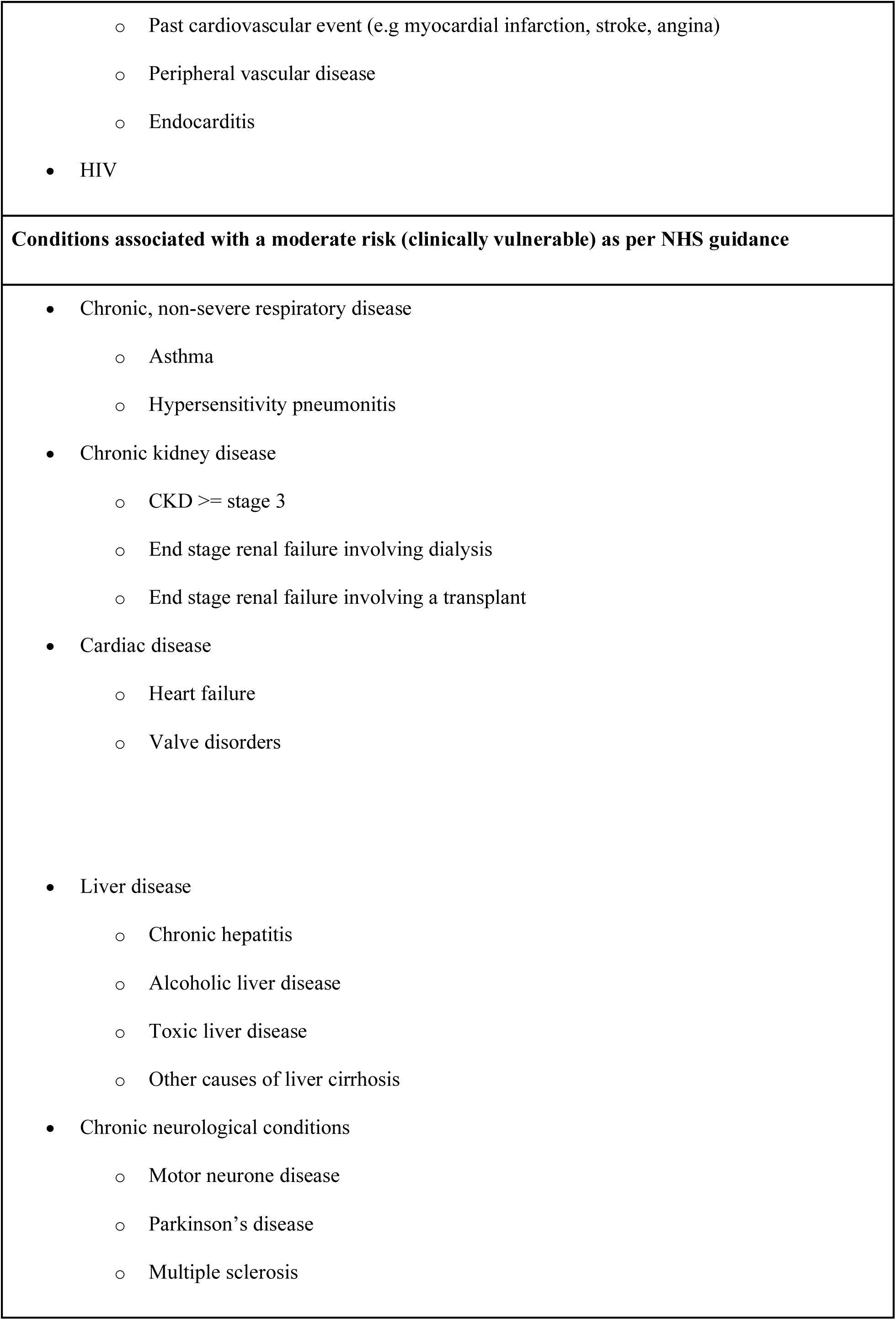
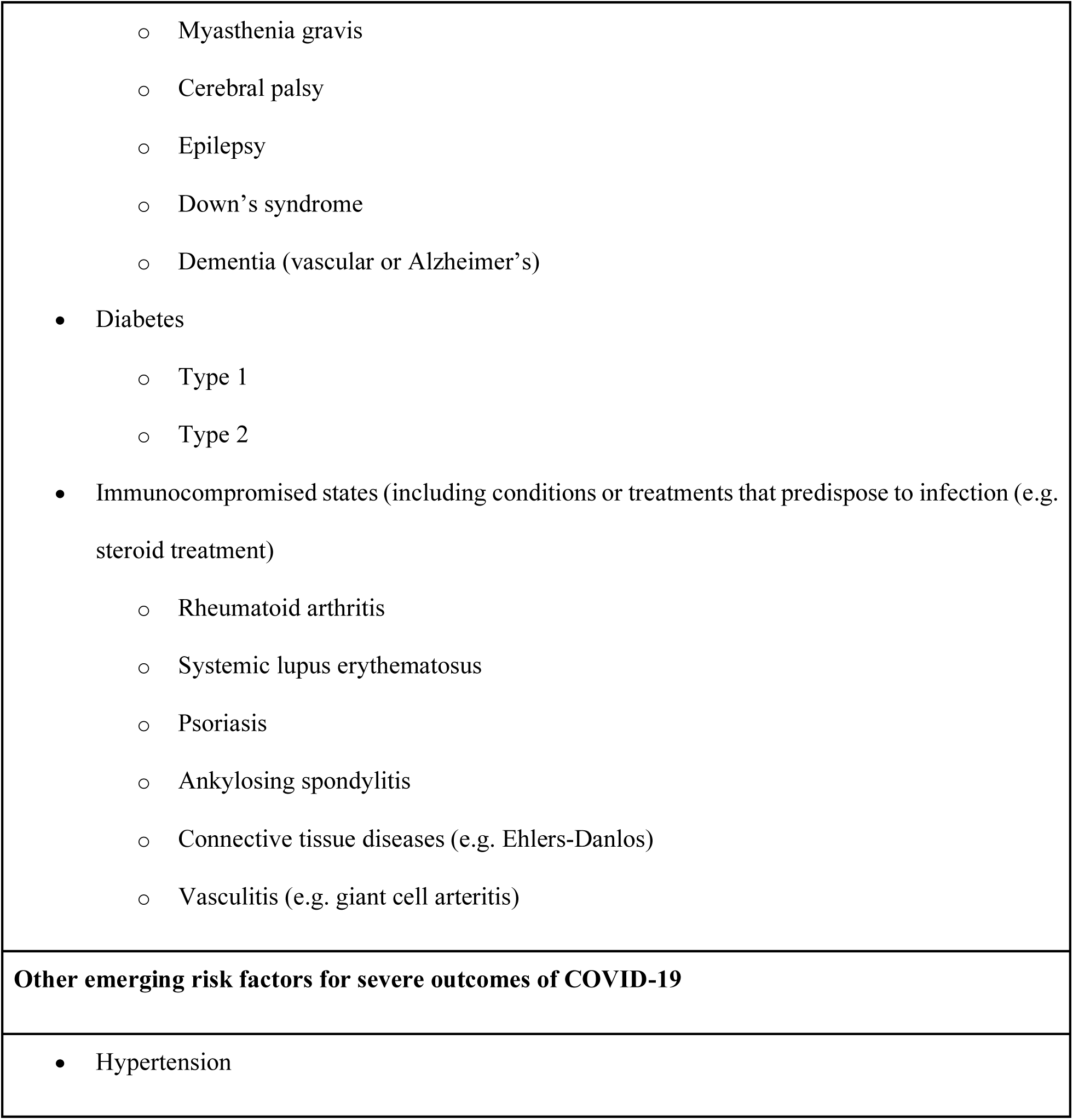
Hand-Crafted Predictor variables

**Figure 2.**
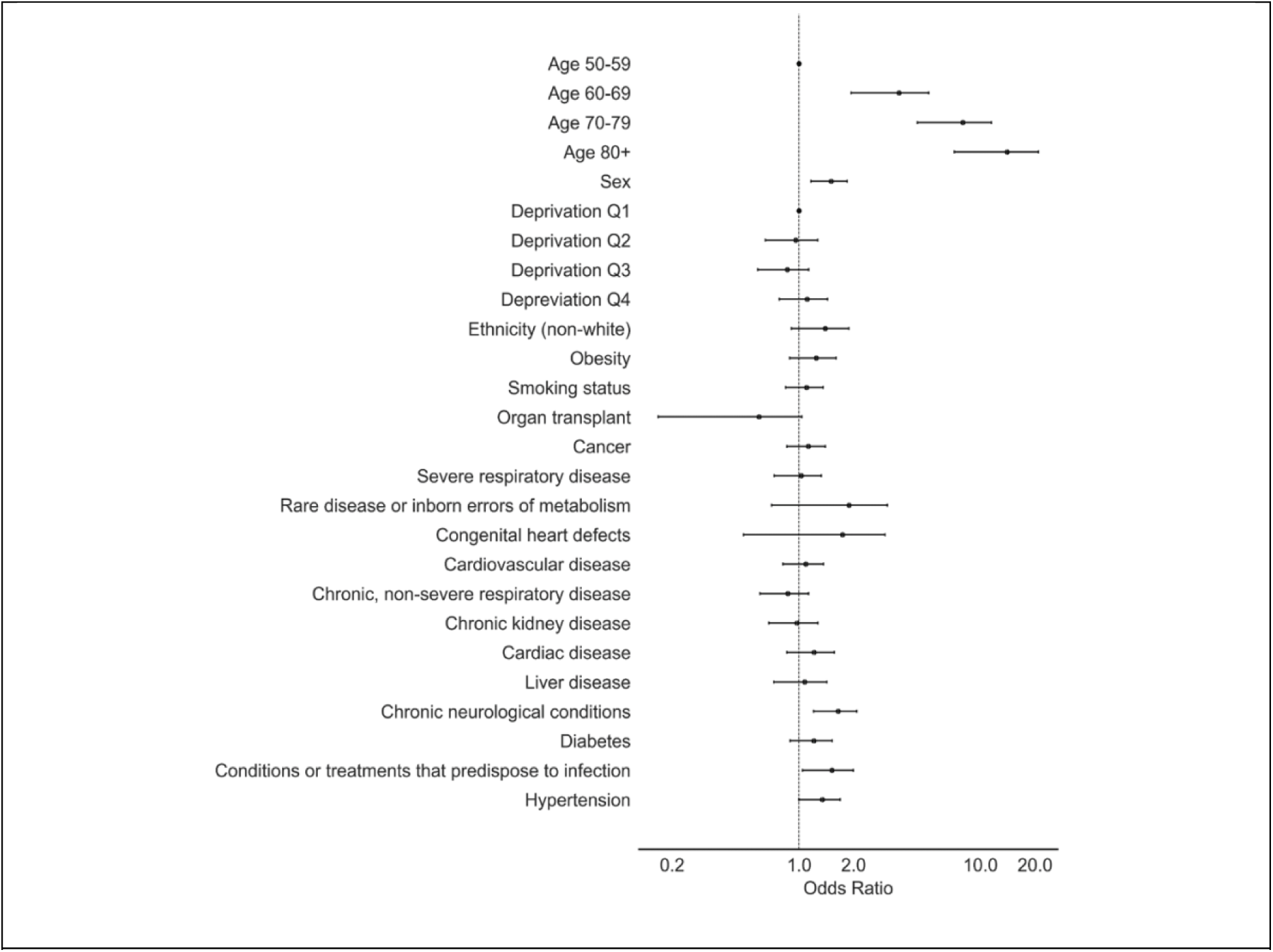
Estimated Odds Ratio for each patient characteristic from a multivariable logistic regression

### Digital Comorbidity fingerprinting

Pre-existing conditions are coded as thousands of possible ICD-10 disease codes in patient records. Our methodology adapts unsupervised methods from Natural Language Processing (NLP) trained on the general population (i.e. the entire UK Biobank which includes over half a million patients) in order to cluster patients’ conditions using only disease codes from a patient’s past hospital visits. We use a form of Latent Dirichlet Allocation, a Bayesian clustering method that can naturally model the categorical nature and inherent imprecision of disease codes. This clustering distils thousands of disease codes, and billions of possible combinations into a set of DCFs. Each patient’s pre-existing conditions are thus summarised by a single number (between 0-100%) describing how strong a specific pre-existing condition fingerprint is present in their record. All pre-existing conditions and their myriad of combinations are summarised by a total of 30 DCFs.

We describe hereafter how a second interpretable machine learning algorithm is used to classify participants with suspected or confirmed COVID-19 into a high and low COVID-19-related death risk category based on their digital precondition fingerprint and their demographics.

We initially fitted our DCF Model on the 402,902 participants who belonged to the UK Biobank cohort, were still alive in January 2020 and were part of the Hospital Episode Statistics (HES) sub-database. Latent Dirichlet Allocation was used to generate the topic distributions for all patients and ICD-10 code group. We tested multiple values of topics (*k* ranging from 2 to 100) and compared the resultant topic distribution with respect to the model coherence, *C*_*v*_^32^. The best results were obtained for *k* = 30 topics, and Dirichlet priors α = 0.1 and η = 0.6. We clinically reviewed disease codes grouped in each of the generated topics, i.e. the DCFs. We observe that our comorbidity fingerprinting does not group codes simply by clinical system (e.g. respiratory disorders or diseases of the circulatory system) as can be seen in Figure 3. Rather, the method groups past clinical codes in a more functional way (all precondition fingerprints are listed in the Supplementary Box 2). For example, *DCF* 24 groups codes of cardiovascular disorders together (such as angina and myocardial infarction), but also includes some of the most common risk factors for these diseases, such as lipidaemia or family history of cardiac disorders. *DCF* 7 groups codes for rhythm disorders (e.g. atrial fibrillation) with pulmonary embolisms. Precondition fingerprint *DCF* 22 groups diabetic disorders with occurrences of renal failure and obesity. Other fingerprints, such as *DCF* 5, *DCF* 8 and *DCF* 19 are mainly defined by neoplasm and surgical procedures (e.g. acquired absence of organs).

**Figure 3.**
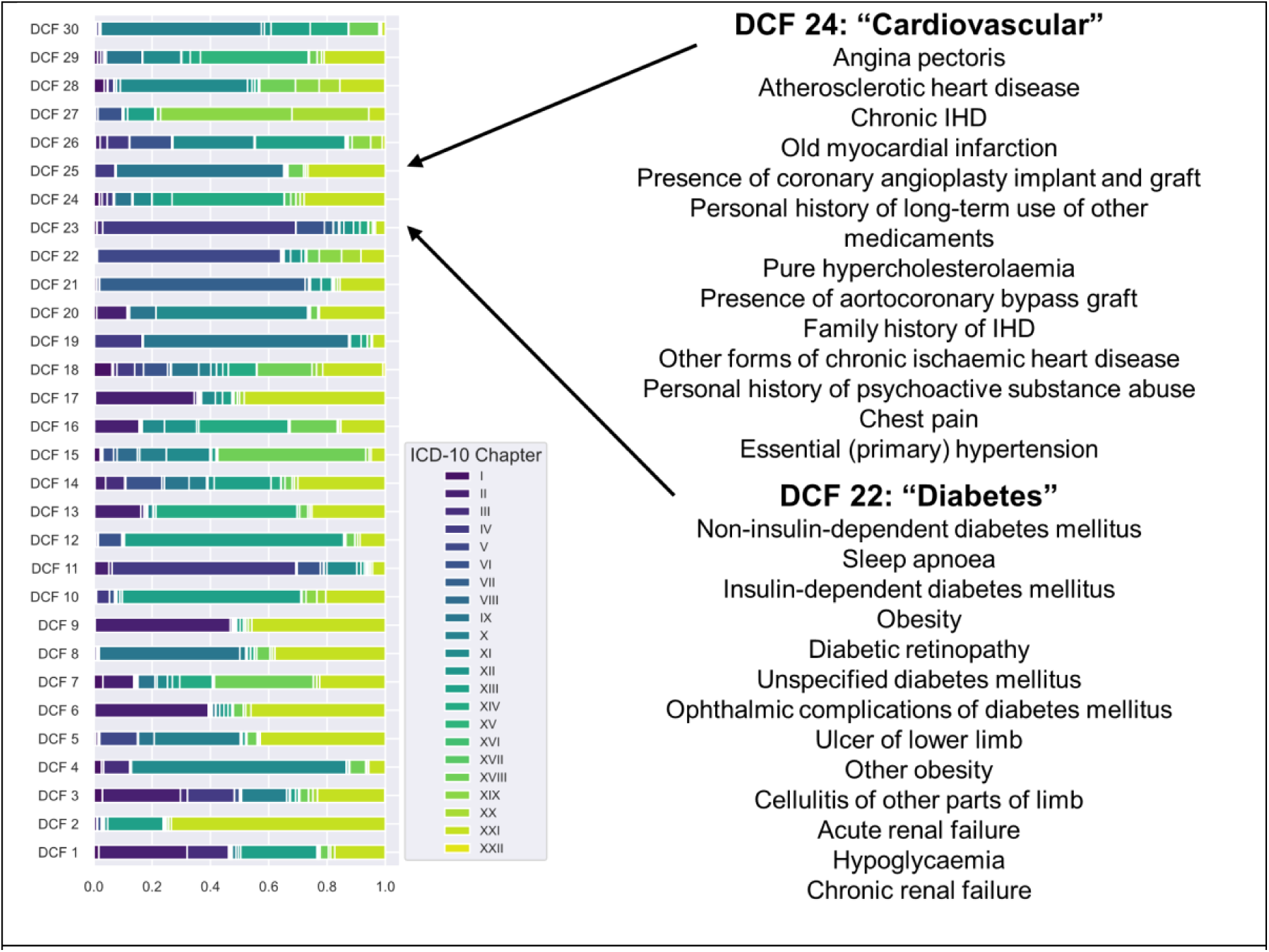
Topic description. We fit a 30-topic LDA model to 420,000 patients from the UK Biobank. On the left is the distribution of the 22 ICD-10 ‘Chapters’ (Supplementary Box 1) over the 30 topics. On the right are the top 13 ICD-10 codes from topics 22 and 24 (in rank order).

These results indicate that comorbidity fingerprinting is able to yield clinically relevant topics by grouping diseases that often appear simultaneously. Some of the clusters are harder to interpret than others and would need to be further investigated. The DCFs are further described in Figure 4, where we demonstrate the ability of the DCFs to differentiate between patients that died of COVID-19 and the overall UK Biobank cohort. Figure 4 further outlines the DCFs ability to encompass notions of gender, age and BMI. It is, nonetheless, clear that comorbidity fingerprinting does generate coherent and interpretable results when applied to disease codes.

**Figure 4.**
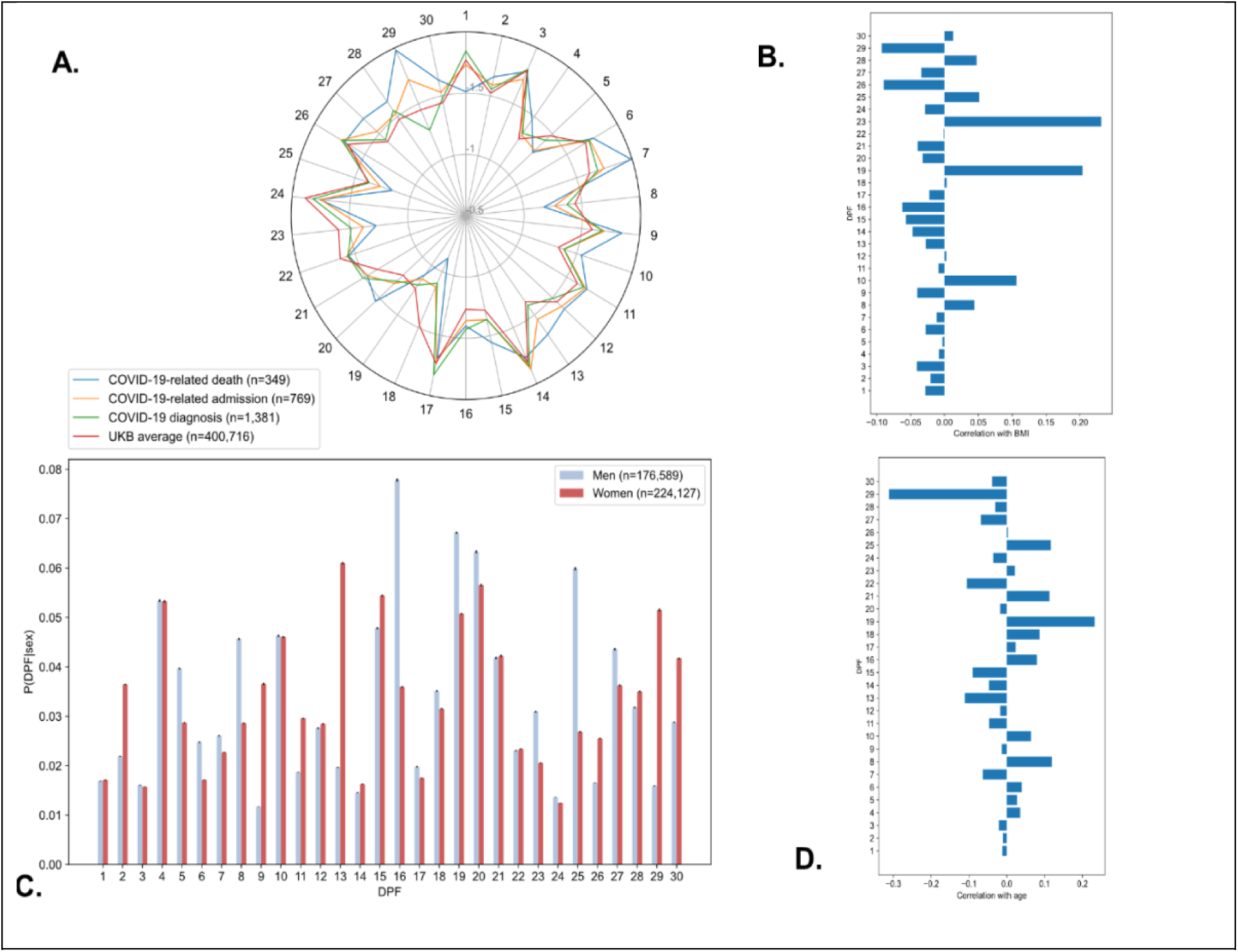
Summary of the Digital Precondition Fingerprints. (A) Details the average probability of belonging to each DPF (log 10 scale) by COVID-19 outcome. (B) Shows the Pearson correlation between each of our DCFs and the patient’s BMI. (C) Shows the average probability of belonging to each DCF by gender and (D) shows the Pearson correlation between the each DCF and the patient’s age.

### Prediction accuracy

In order to evaluate our DCF approach to feature engineering, we used it as an input to a supervised classifier of COVID-19 mortality and assessed its discrimination capabilities when compared to a model built using the EHCF. The prediction accuracy for the two models under consideration are shown in Table 2 and Figure 5. We use a model built from age alone as a baseline for performance evaluation (area under the receiver operating curve (AUC-ROC): 0.709 95% CI: 0.692 - 0.727). Both the EHCF Model, (AUC-ROC: 0.734, 95% CI: 0.704 - 0.764) and the DCF Model, (AUC-ROC: 0.730, 95% CI: 0.700 −0.760) achieved similar discrimination scores. A Wilcoxon’s rank-sum test was performed to assess if our DCF Model showed better discrimination capabilities than the baseline. Although there was a small absolute difference between these two models (a difference of 0.021), the *p*-value was significant at 0.0409.

**Figure 5.**
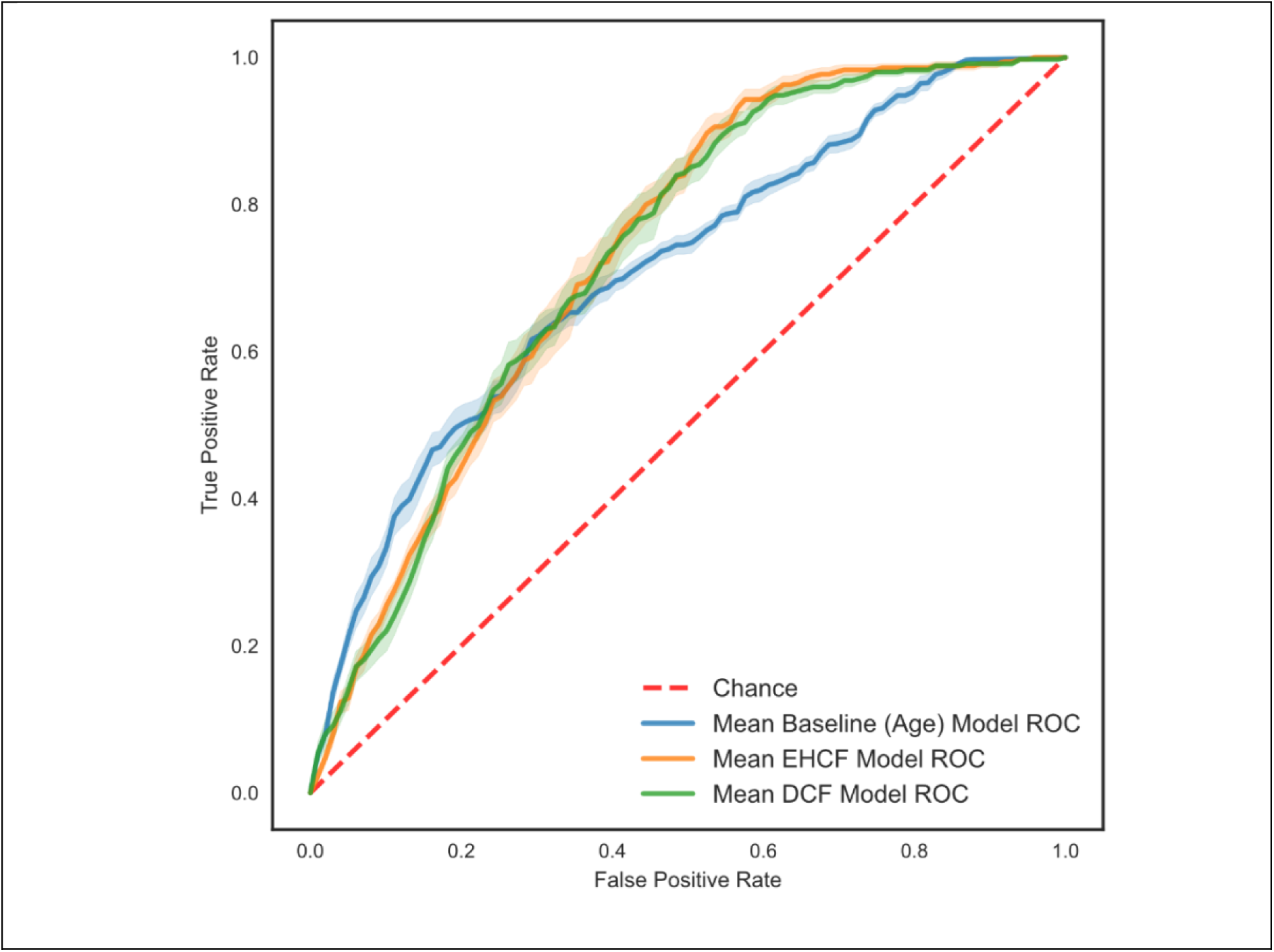
Mean receiver operating characteristic curves derived from predicting COVID-19-related death in the 10-Fold Cross Validation (standard error of the mean) for all models.

**Table 2.** Performance of all prediction models under consideration

| Predictor Set | Results |  |
| --- | --- | --- |
|  | AUC-ROC | 95% CI |
| Baseline | 0.709 | 0.692 - 0.727 |
| EHCF - Model | 0.734 | 0.704 - 0.764 |
| DCF - Model | 0.730 | 0.700 - 0.760 |

### Model interpretability and impact

Comparing the EHCF Model to our DCF Model showed overlap in the top contributing factors for predicting COVID-19-related death (Figure 6). Age is the feature that plays the most predominant role in both models. Nine of the other top ten ranked predictor variables (diabetes, renal disease, hypertension, respiratory diseases, sex, predisposition to infection and cardiovascular diseases) were identified in both models. Indeed, the EHCF Model highlights the importance of cardiovascular diseases, diabetes, sex, cardiac diseases, severe respiratory disease, chronic kidney disease, conditions that predispose to infection and cancer. These are mirrored in *DCF* 24 (cardiovascular diseases), fingerprint *DCF* 22, (severe diabetes and obesity), fingerprint *DCF* 4 (chronic pulmonary disease and tobacco usage), *DCF* 18 (chronic kidney failure and hypertension), *DCF* 0 (haematological disorders and immunosuppression). *DCF* 8 in the DCF Model reflects some preconditions of the female sex (acting as an indicator for the sex variable, see Figure 6) and some associated diseases by concentrating on female-specific cancer codes such as breast cancer and more generic ICD-10 cancer codes such as chemotherapy sessions. Chronic neurological conditions are important predictors in the EHCF Model but this was not directly identified as important in the DCF Model. Instead, the DCF Model puts emphasis on gastrointestinal cancers and disorders of the GI tract through the importance of *DCF* 19, this is a feature not covered in the EHCF Model. Thus, we obtained a different set of features in our comorbidity fingerprinting (DCF), as it encompassed significantly more comorbidities per topic, than feature sets assembled by-hand (EHCF).

**Figure 6.**
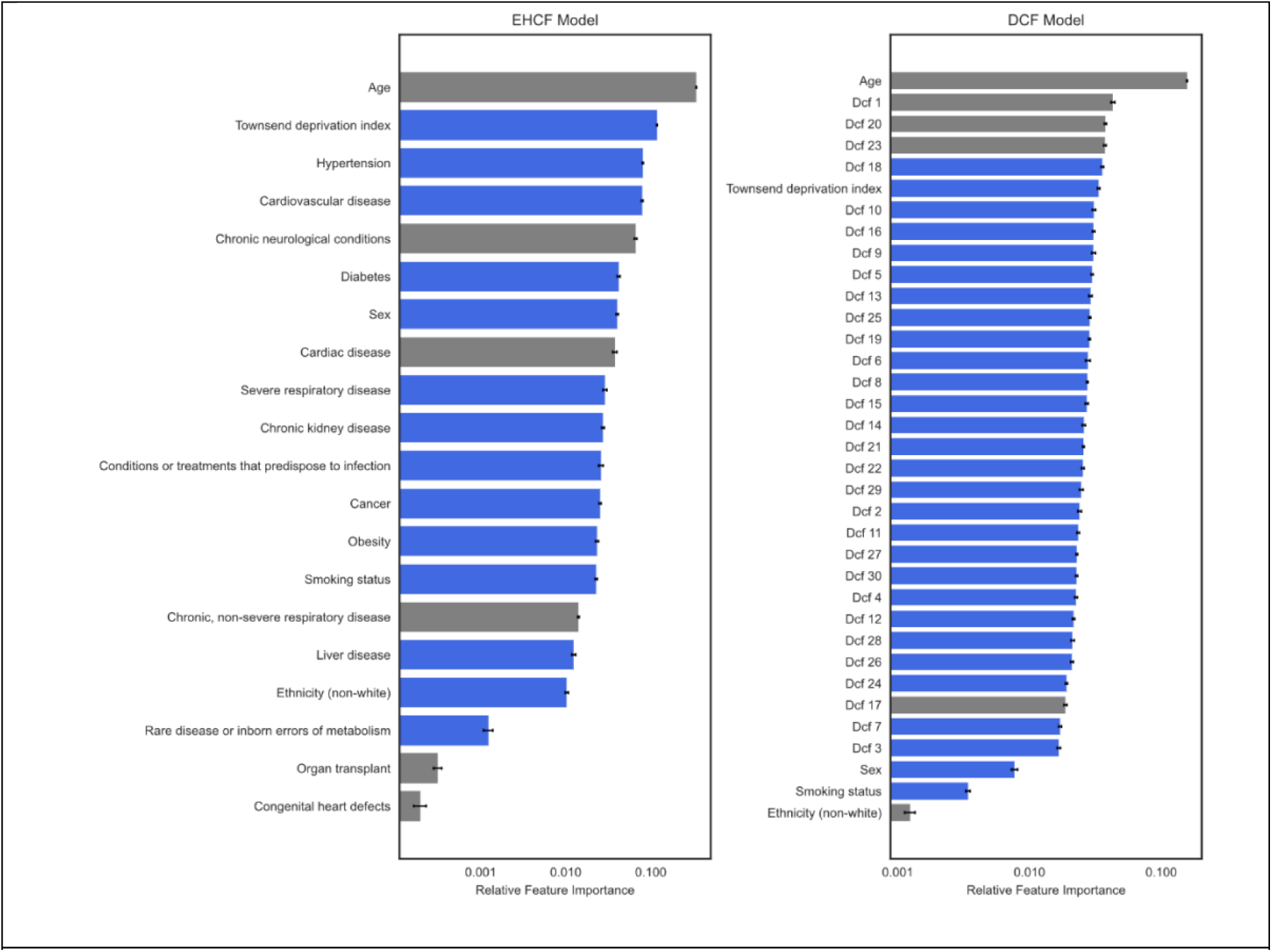
Comparison of relative feature importance (note logarithmic scale) showing the contribution to the predictions of the EHCF and our DCF Model. In grey the demographic and lifestyle features which were the same in both models and in blue the model specific features. Note, how in the DCF Model aside from age the co-morbidity and multi-morbidity make up the key 15 features.

## Discussion

Our findings demonstrate how taking a strictly data driven approach using disease codes directly from EHRs can be rapidly leveraged for predicting outcomes and studying disease patterns. The model developed in this study required, in addition to the previously available data (prior to January 2020), only diagnosis information and cause of death, to predict who, with a positive COVID-19 test, was likely to die. Even though our model does not rely on symptoms, laboratory values or images (at the time of diagnosis or during the illness) to predict mortality, we are able to achieve comparably high discrimination results (ROC-AUC of 0.703). Other published COVID-19 prognosis models have AUC-ROC’s ranging from 0.68 to 0.90^33–35^. The use of physiological data at the point of admission further makes these models difficult to implement in a community setting or use a specific predefined subset of comorbidities^36^.

In order to evaluate the capabilities of our topic model-based approach to clinical dimensionality reduction, we compared it to current medical risk prediction approaches ^19, 35^. We opted to develop a set of expert hand-crafted features, built using the CDC and NHS COVID-19 shielding list combined with current research. Our method showed comparable discrimination capabilities, (ROC-AUC of 0.730) to the current expert models (ROC-AUC of 0.734). The significance of our AI-based methodology is highlighted by the ability of our model to encompass the entire past medical history of the participants and, in the context of a new and unknown disease, not be restricted by potentially biased prior belief. In this way, the automated, data-driven approach we present here is able to avoid human-induction bias in the classifier’s input features and permits the detection of novel insights in a new disease in a very short amount of time and with minimal human supervision. Furthermore, by ensuring a fully automated process, the methodology we present here is easily scalable to other healthcare systems and generalisable to future events. Furthermore, the DCFs we present here are not specifically tied to COVID-19 risk prediction and can thus be immediately reused to summarise risk predictions for COVID-19 variants or other diseases.

Our study provides results regarding key findings in the underpinning structure of comorbidities and validates the usefulness of these associations with their predictability of COVID-19 mortality. This is of particular importance given the high UK prevalence (18.5 million individuals) of underlying conditions that increase the risk of severe COVID-19^37^. Our DCFs offer a rich and deep set of features, that can succinctly summarise a patient’s comorbidity profile using topic modelling. This approach allows for the use of a high-performance AI algorithm whilst remaining clinically interpretable and intuitive. Some obvious common causes of comorbidity clusters are well known (e.g. cardiovascular disease due to smoking or obesity^38^), advances in the science of multi-morbidities are challenging because diseases may have linked or independent causes, combine heterogeneously, are treated differently, and thus affect care pathways and medication strategies. Structuring the myriad of combinations of comorbidities, such as through our comorbidity fingerprinting, underpins any measures to slow the accumulation of conditions and to optimise their treatment. We have shown that AI methods can help in identifying multimorbidity clusters without human-induction bias, while at the same time ensure that findings remain clinically useful.

We further demonstrate that the DCFs generated using LDA correspond well and show significant variable importance overlap when compared with the EHCF Model and previously identified risk factors from published epidemiological studies. For example, our model supports the previous epidemiological findings that cardiovascular disorders and hypertension increase risk of COVID-19-related death ^12, 13, 19^. Several features identified in the model warrant further evaluation as they were not specifically seen in the initial COVID-19 epidemiological studies. DCF 19 ‘GI Disorders’ was one of the most important features in our model, however many of the epidemiological studies do not report this as a significant risk factor ^13, 31^.

There are a few limitations to our study. Firstly, our data set while vast, may also reflect the inherent bias of the UK Biobank^39^, which has been discussed in detail elsewhere^39–41^; notably, the demographic reflect a “healthy volunteer” bias, with individuals being generally older, from more educated, less deprived socioeconomic backgrounds, and with significant under-representation of ethnic minorities compared to the UK population. Secondly, testing, treatments, and outcomes of COVID-19 have continuously improved during the study period, thus possibly having a confounding effect on the results. Moreover, due to the limited availability of testing kits for COVID-19, priority was initially offered to those considered at a higher clinical risk, thus potentially leading to an overestimation of severe outcomes in the database. However, these COVID-19 specific data bias factors only affect the mortality prediction but not the structure of the DCFs. This study further relied on retrospective secondary care EHRs and the model is therefore currently blind to conditions entirely managed in primary care, such conditions include many less severe cases of diabetes, asthma and hypertension. At the time of the study, data from primary care was not available. Future work will be needed to incorporate data from General Practices into the development of the DCFs. Balancing the strengths and limitations, we consider our derivation population to be relevant for the initial exploration of COVID-19 mortality in an older and more ‘at risk’ population using a machine leaning approach. Further research and external validation of the predictive models and DCFs in this study will be required before applying this work in a more general context, specifically to better asses the risk in a younger and fully diverse population.

In conclusion, we developed a novel machine learning based approach to predict COVID-19 mortality in a community cohort only from past EHR data. We have demonstrated the feasibility of a comorbidity clustering approach to avoid human-induction bias and succinctly summarise billions of possible comorbidity combinations in COVID-19 prognosis modelling and thus rapidly leverage vast EHR dataset for outcome prediction. We find, that age and qualitative co-morbidity information are very powerful mortality predictors, making other demographic or physiological data pale in comparison. This implies, that in rapid decision situations or decisions with limited information will benefit from our solution as this enables healthcare professionals to form a rapid picture from information that patients and their relatives are familiar with and which do not change rapidly, unlike data such as blood pressure. Thus, our model may enable early stratification of key clinical risk groups thus permitting earlier intervention and may help better prioritise risk groups in the global vaccination effort.

Crucially, however, our DCF framework can be applied to any other form of novel disease as the multi-morbidity features we discovered span a space that is disease agnostic.

## Methods

### Study design and participants

We conducted a prospective open cohort study using the Hospital Episode Statistics for England (HES) database, COVID-19 test results and death data linked through the UK Biobank (see ‘Data Source’). The cohort study began on 30 January 2020, which was chosen as it corresponds to the day of the first laboratory-confirmed case of COVID-19 in the UK^42^; and ended on 9 September 2020. The cohort study examines risk among the population of patients who are positive for COVID-19. Therefore, we only included patients from the UK Biobank if they had a positive SARS-COV-2 test result or a COVID-19 diagnosis code recorded in their clinical notes (ICD-10 code U071 or U072).

### Data source and ethical approval

The UK Biobank is a large prospective population cohort of 502,505 participants aged 40-70 years at baseline, prepared to travel to 1 of 22 assessment centres in England, Scotland, and Wales^23^. All participants were recruited between 2006 and 2010, and all consented to have their health followed^23^. Baseline assessments include nurse-led interviews surrounding socio-demographics, lifestyle, physiological measurements, and medical history. Health outcomes were sourced from linkages to electronic medical records. UK Biobank obtained approval from the North West Multi-Centre Research Ethics Committee (MREC), and the Community Health Index Advisory Group (CHIAG). All participants provided written informed consent prior to enrolment in the study. The UK Biobank protocol is available online^43^.

In the context of the COVID-19 pandemic, the UK Biobank has implemented a rapid and dynamic linkage between the laboratory results for COVID-19, stored on Public Health England’s Second-Generation Surveillance System, and the UK Biobank participants^24^. The previous hospital medical information of the study participants was obtained from linkage to the HES database (a data warehouse containing records of all admissions, emergency room attendances and outpatients appointments at NHS hospitals in England)^44^.

Access to anonymised data for the UK Biobank cohort was granted by the UK Biobank Access Management Team (application number 21770). Ethical approval was granted by the national research ethics committee (REC 16/NW/0274) for the overall UK Biobank cohort.

### Outcome

The primary outcome measure was COVID-19-related death in patients with a COVID-19 infection. Confirmation of a COVID-19-related death COVID-19 death was defined as 1) a COVID-19-related death recorded in ONS death certificate data (International Classification of Disease, 10th edition, ICD-10 codes U071 or U072). 2) a death occurring within 28 days of a laboratory-confirmed COVID-19 infection (as per UK government guidelines^45^).

### Model Development

The performance of machine learning methods is heavily dependent on the selection of features^46^. For that reason, most of the effort in an analytic model is spent pre-processing, merging, customizing, and cleaning datasets. This task is made all the more complicated in EHRs as the different predictors may number the tens of thousands (over 11,700 are present in the UK Biobank). Traditional modelling approaches deal with this complexity by choosing a limited number of variables and creating custom features^47^. However, this is a largely manual and often labour-intensive task. Another common approach is to use unsupervised dimensionality reduction approaches, this is often far faster but may yield results that are less accurate or interpretable.

### Hand-Crafted features: Predictor variables

The patient-level characteristics included in the development of the EHCF Model were selected from the existing health conditions listed on original population-level risk stratification method as exercised in UK^17^ or listed by the Centers for Disease Control and Prevention (CDC) as defining ‘people at increased risk for severe illness’^18^. This was further complemented with other emerging risk factors for severe outcomes of COVID-19 (such as raised blood pressure)^9, 12, 13, 19^. The final predictor variables chosen are summarised in Box 1.

Demographic and lifestyle variables were also included, namely: age, sex, Townsend deprivation index^48^, ethnicity, and smoking status. Age groups were categorised as 50-60, 60-70, 70-80 and 80+ years. Deprivation was measured using the Townsend Deprivation Index (grouped into quartiles based on the entire UK Biobank distribution, greater scores imply a greater degree of deprivation). Ethnicity was grouped as white and non-white (thus collapsing the UK Biobank categories mixed, asian, black or chinese), due to the otherwise small number of participants in each of the specific categories. Smoking status was determined from data from the initial UK Biobank assessment and grouped into participants that smoke or have smoked (current or previous at baseline) and participants that, at baseline, were never smokers.

The demographic variables considered as potential factors affecting COVID-19 risk and were determined using the answers given at UK Biobank initial assessment centre. Age was determined on 30 January 2020. Information on all other covariate variables was determined using the ICD-10 codes in the linked HES database.

### Automated features: topic modelling

Topic modelling was originally developed as a tool to model collections of discrete data with particular application in text modelling and Natural Language Processing (NLP). Topic modelling can discover abstract ‘topics’ within a collection of documents, where a topic consists of a collection of words that frequently occur together. Here we opted to train topic models using Latent Dirichlet Allocation (LDA), an unsupervised and interpretable method for topic modelling^49, 50^. LDA is a Bayesian probabilistic model that works by taking as input a corpus of clinical codes and represents each document as a finite mixture of an underlying set of fixed topics, with each topic characterised by its distribution over words^49, 51^.

Here, LDA is used to represent a patient *p* as a mixture over the collection of *K* “topics”. Each topic *k* defines a multinomial distribution over a finite vocabulary of ICD-10 diagnosis codes and is assumed to have been drawn from a Dirichlet, β_*k*_ ∼ *D*irichlet(η). Thus, each code *c*_*p*,*n*_ has a given probability *β*_*k*_ in each topic *k*. Given the topics, LDA then generates for each patient *p* a distribution over topics θ_*p*_ ∼ *D*irichlet(α). In other words, each topic is determined as a distribution over closely related codes and each patient is represented as a distribution over multiple topics. For simplicity, we assumed symmetric priors on the patient topic distribution *θ* and the code topic distribution *β*.

For each participant of the UK Biobank, we considered a full longitudinal visit history to hospital, including all admissions, emergency room attendances and outpatient appointments. For each recorded hospital episode, we extracted a list of diagnosis codes, using the ICD-10. This generated a document for each unique patient in the UK Biobank, representing their full hospital visit history as a series of ‘sentences’ (one for each visit, so if a patient visit the hospital 10 times for asthma it is represented 10 times in their record.), wherein each ‘word’ was a unique ICD-10 diagnosis code. Thus, this way of accounting naturally weighs conditions that require more frequent visits to the hospital.

The LDA topic model was trained using as input the corpus of documents (i.e. sentences of ICD-10 diagnosis codes) for the entirety of the UK Biobank population that is not lost to follow up, still alive on 30 January 2020 and is part of the HES database (‘Population part of the HES database’, Fig. 1). This was chosen with the aim of generating more clinically robust and disease agnostic topics (i.e. the topics formed are not specific to any prior comorbidity). From this corpus, we used a grid-searching method to determine the optimal number *K* of topics, and the prior value for θ and β (α and η) when optimising with respect to model coherence, *C*_*v*_ in^32^ as this method has been shown to achieve the highest correlation with human topic ranking data^32, 52^. This model was implemented using the gensim library in the Python programming language^53^.

### Development of a random forest model

Two predictive classifiers were trained in this study, EHCF Model and DCF Model. The first, EHCF Model, uses as input the ‘manually’ determined participant characteristics or ‘Hand-Crafted Features’ (see Hand-Crafted features: Predictor variables). These were grouped into the aforementioned categories and each participant was set a binary value indicating whether or not their past medical records (extracted from the HES database) showed the occurrence of at least one of the conditions for each category. The DCF Model, on the other hand, uses the patients’ distribution over the previously derived topics of an LDA topic model as input to a classifier. The LDA model was further complemented with the demographic and lifestyle variables (e.g. age).

The models described above were trained to predict the same binary output; the patients that survived were assigned to class 0 and those that died to class 1.

In this study, we opted for use of a random forest (RF) classifier^54^, to determine COVID-19 associated mortality. RF is a high-performance ML algorithm that fits several decision tree classifiers on various sub-samples of the dataset and, subsequently, averages these tree predictors^54^. This allows for a reduction in over-fitting and improvement in accuracy. We implemented all RF models using the Scikit-lean library in python programming language^55^. The models’ hyper-parameters^56–59^ were determined via grid search with AUC as the response as it yields a finer evaluation than commonly used error rates^58^.

The overview of the pipeline used for DCF Model is illustrated in Figure 7. Figure 7 describes the learning of topics from the data and shows how we use the distribution of each patient over each of these topics as input to a RF classifier.

**Figure 7.**
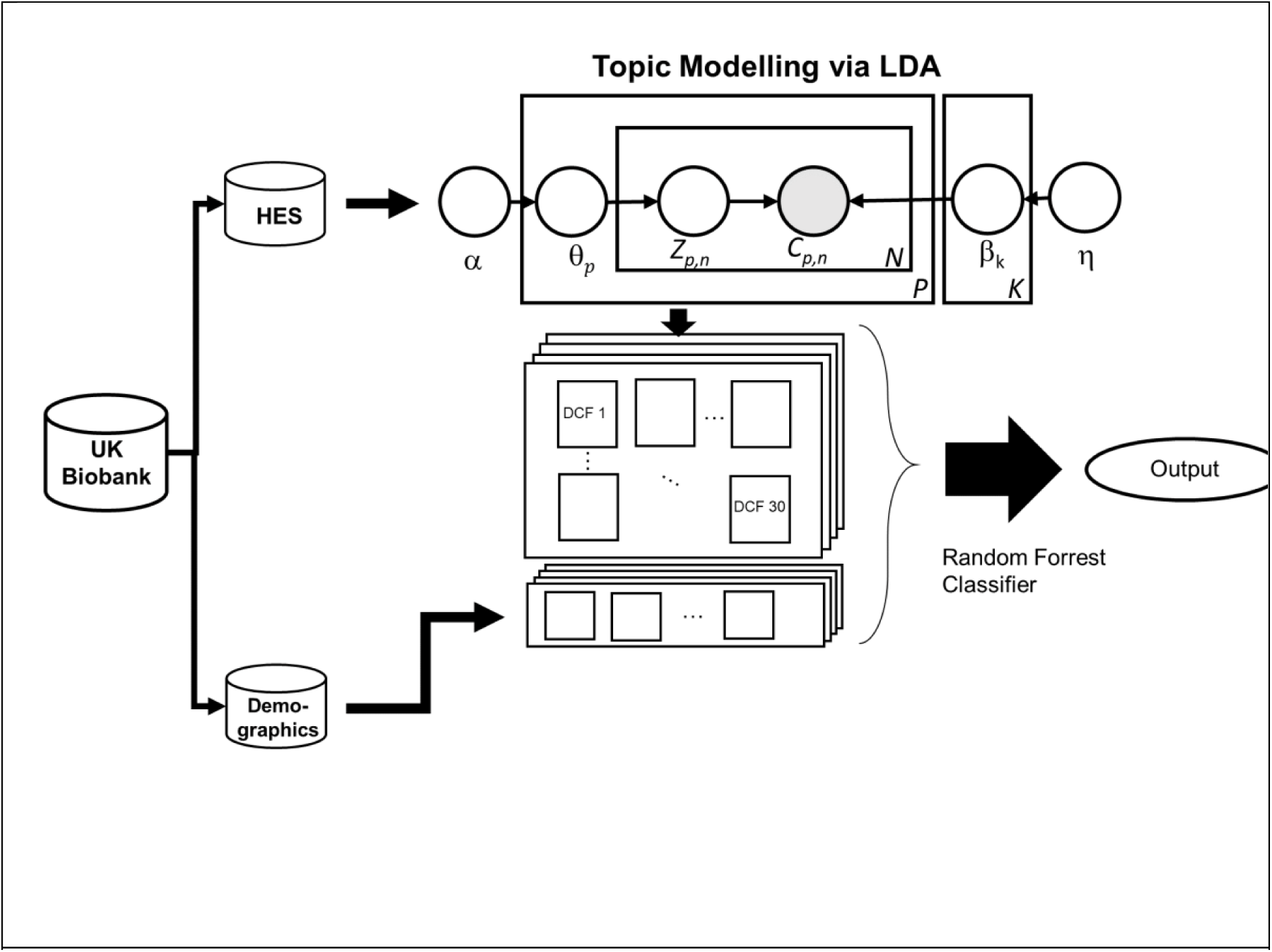
Illustration of the overall study design for the DCF Model. First, we separate the UK Biobank into HES and demographic data, we generate the features (using LDA) from the discrete input of the HES database. We subsequently use the distribution of each patient over these features as input to a supervised random forest classifier to predict COVID-19 fatality.

### Variable ranking

In order to better understand the predictor variables that may lead to increased risk of a COVID-19-related death and their interactions, we use an impurity-based approach to rank the contribution of the different variables in the prediction made by the model. This resulting variable importance score reflects the relative impact that each variable has on the prediction issued by the model.

We chose to use a RF algorithm for this analysis because it is a non-parametric algorithm that can recognise complex patterns and automatically capture nonlinear and interaction effects without specifying these a priori^60^.

### Statistical analysis

Study participant numbers are depicted in the flow chart (Figure 1). For each of the ‘Hand-Crafted’ patient characteristics (see Hand-Crafted features: Predictor variables), a logistic regression model was fitted with the COVID-19-related death as the outcome in order to determine a univariate (not-adjusted) Odds Ratio (OR). All patients’ characteristics were subsequently included in a single multivariable logistic regression to determine the adjusted OR. All OR from the univariable and multivariable logistic regression models are reported with 95% confidence intervals. For the purpose of this study, the logistic regression model was not considered for its predictive power but rather for its ability to simply and accurately describe the available data.

The HES data were considered complete with no missing data for our population. Participants with missing smoking information were assumed to be non-smokers. Missing deprivation score was imputed using the median value of the entire UK Biobank cohort. The participants with missing ethnicity were dropped from the study as no reliable way could be determined to input this field.

We subsequently developed RF based models in order to assess discrimination capability and compare variable importance. In order to avoid over-fitting, we evaluated the prediction accuracy of these models using 10-fold stratified cross-validation and calculated the area under the AUC-ROC as a measure of model discrimination. In every cross-validation fold, a training sample (90% of the participants) was used to derive the models, and then a hold-out sample (10% of the participants) was used for performance evaluation. We report the mean AUC-ROC and the 95% confidence intervals for all models. In all RF models, age and deprivation score are considered as continuous variables.

## Data Availability

The UK Biobank cohort data that supports the findings of this study is available to researchers as approved by the Biobank Access Management Team.

## Data Availability

Data available from central access-controlled repository, and sufficient details included to identify specific dataset. All bona fide researchers can apply to use the UK Biobank resource for health-related research that is in the public interest." This research has been conducted using the UK Biobank Resource under application number 21770.

https://www.ukbiobank.ac.uk/

## Acknowledgements

We are grateful to UK Biobank participants. This research has been conducted using the UK Biobank Resource under Application Number 21770. Infrastructure support for this research was provided by the Imperial NIHR Biomedical Research Centre. AAF acknowledges his UKRI Turing AI Fellowship (EP/V025449/1). The acknowledged parties and funders had no role in the design of the study.

## Author Contributions

Conception and design: ELL, SH, AAF; data analysis: ELL, SH, BK, AAF; data interpretation: ELL, BP, SH, SJB, AAF; writing and revising the paper: ELL, BP, SH, SJB, BK, AAF

## Supplementary Material

**Supplementary Box 1.**
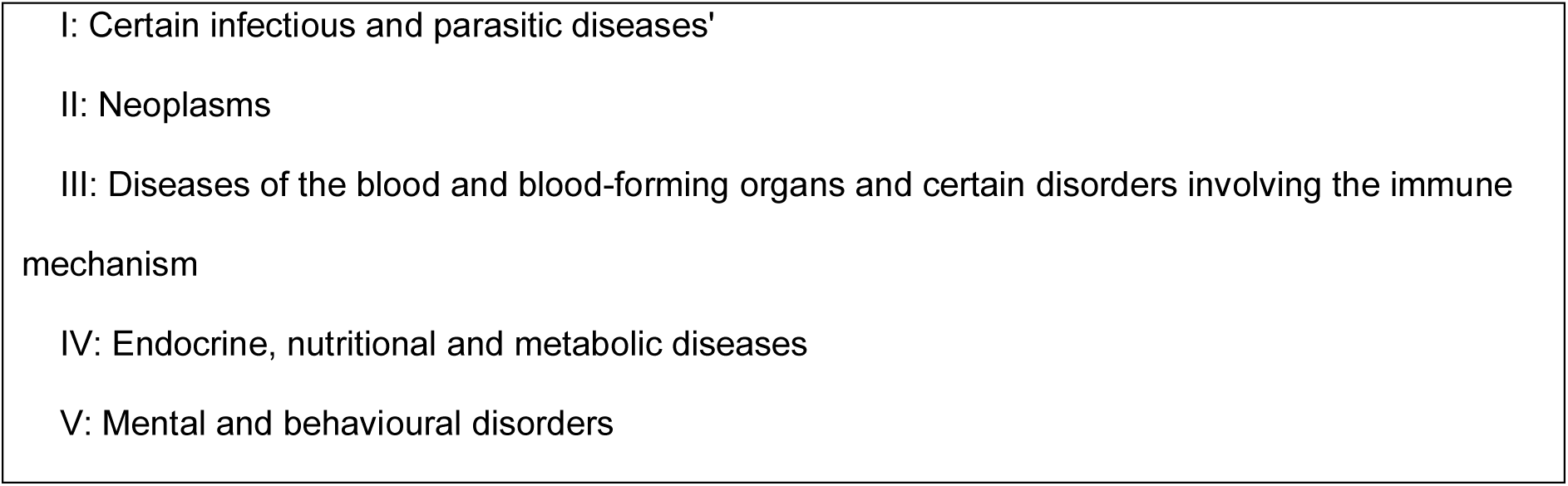
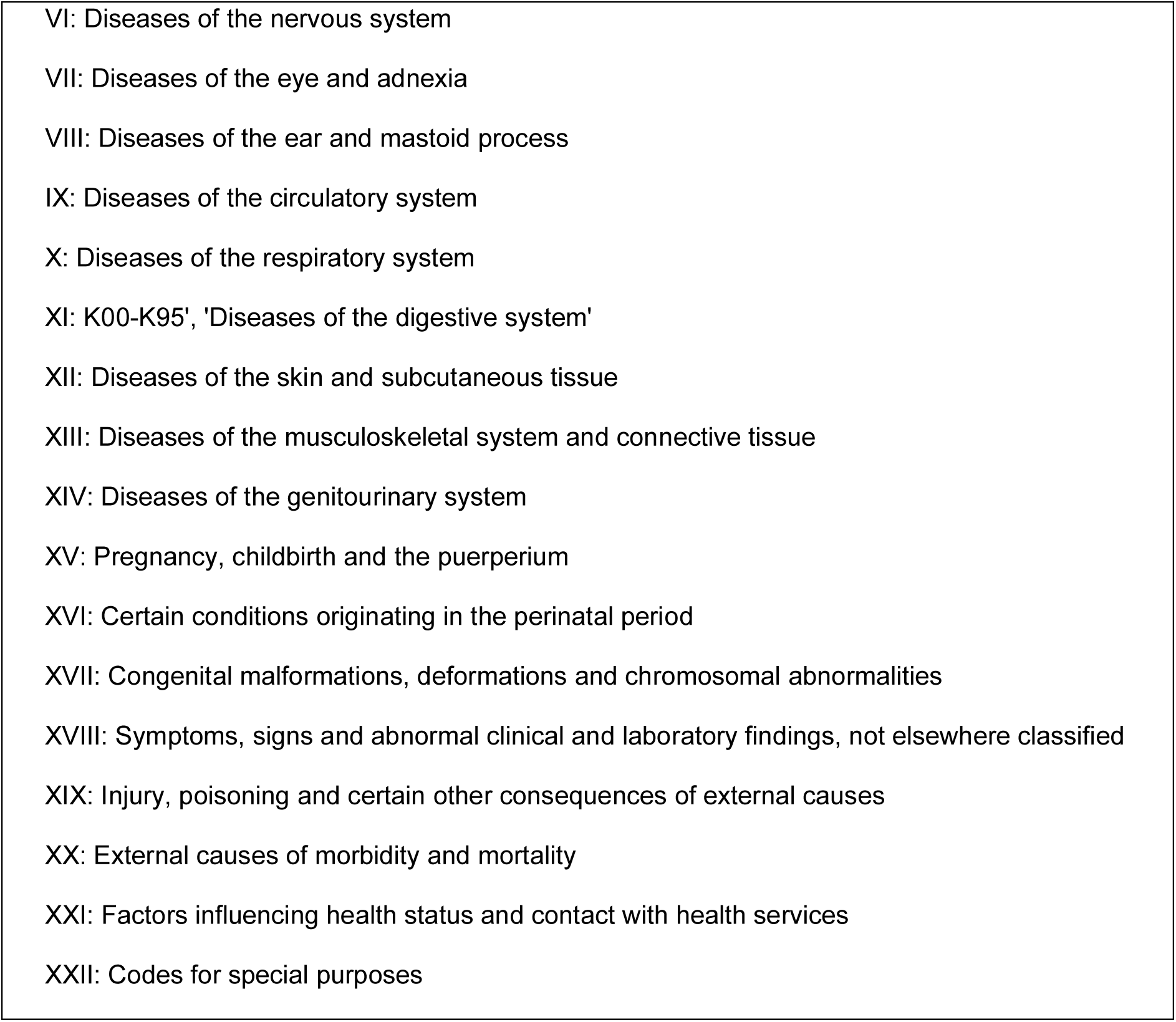
Chapter number and description for each of the 22 chapter’s making up the ICD-10 clinical codes

**Supplementary Box 2.**
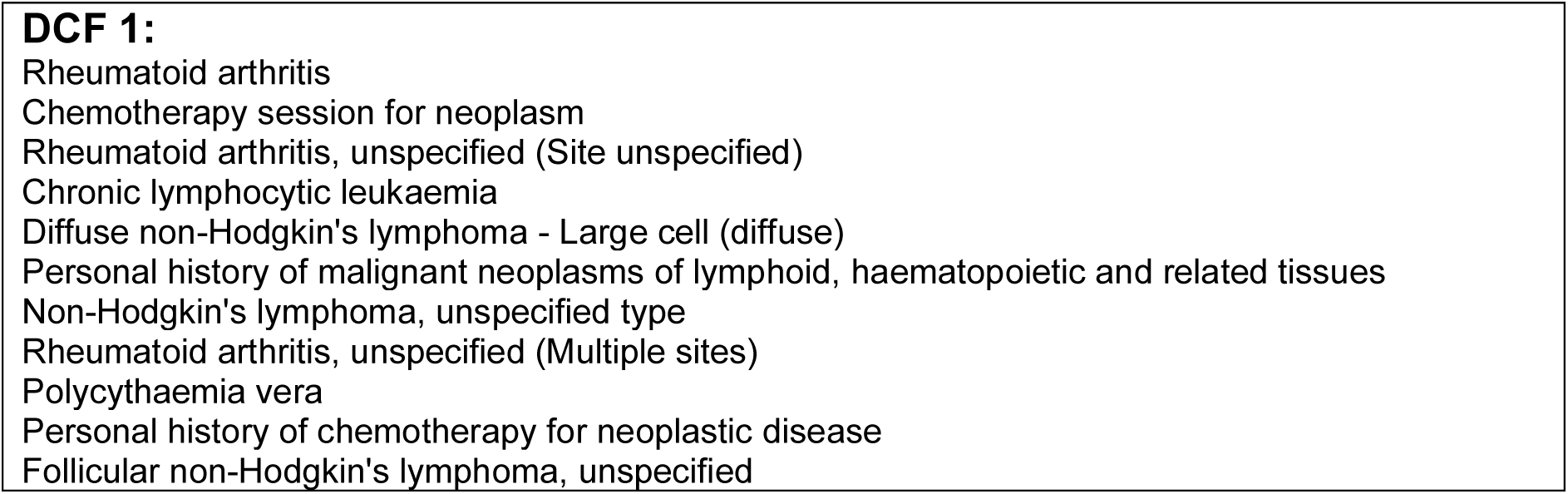
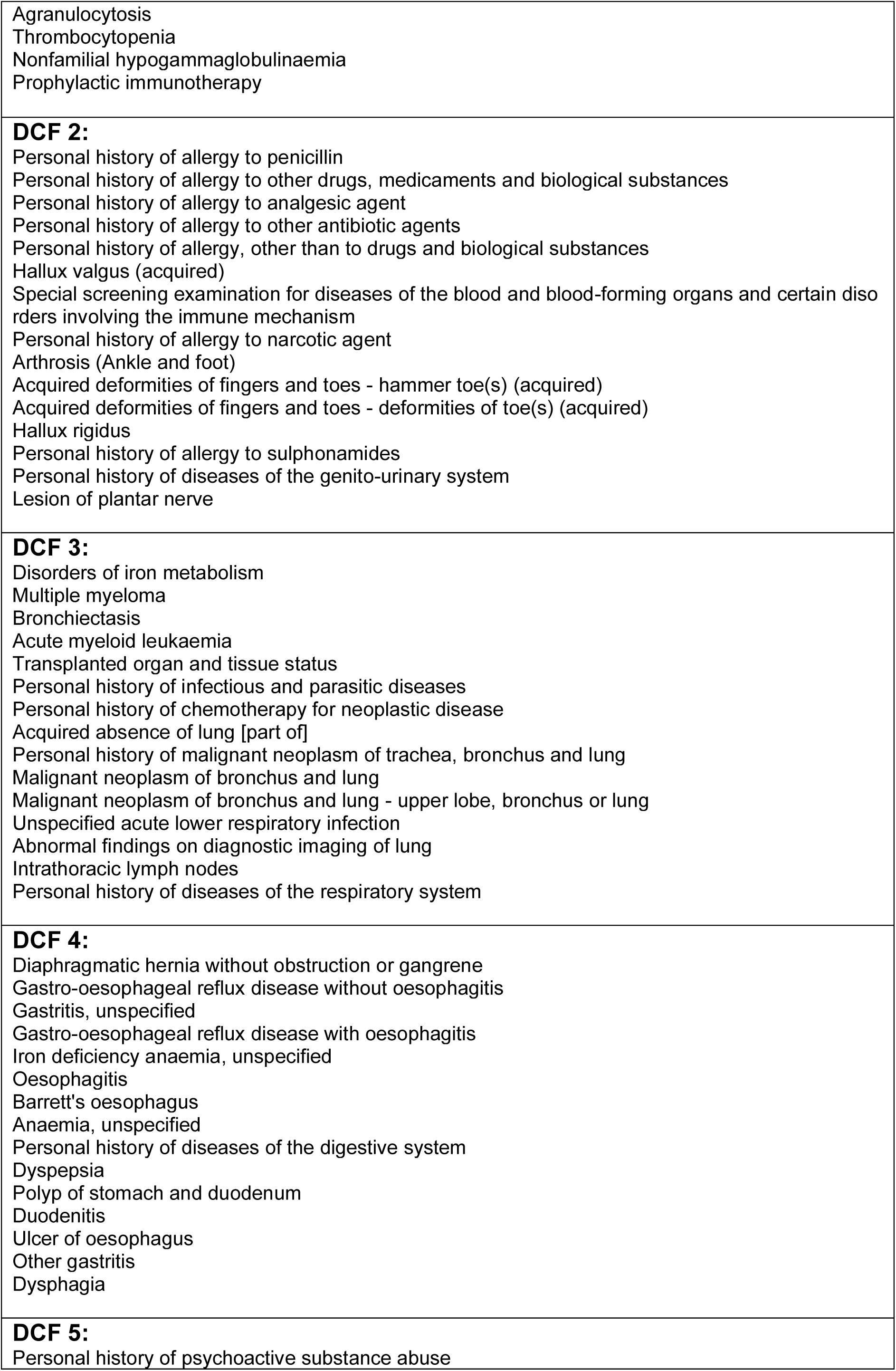
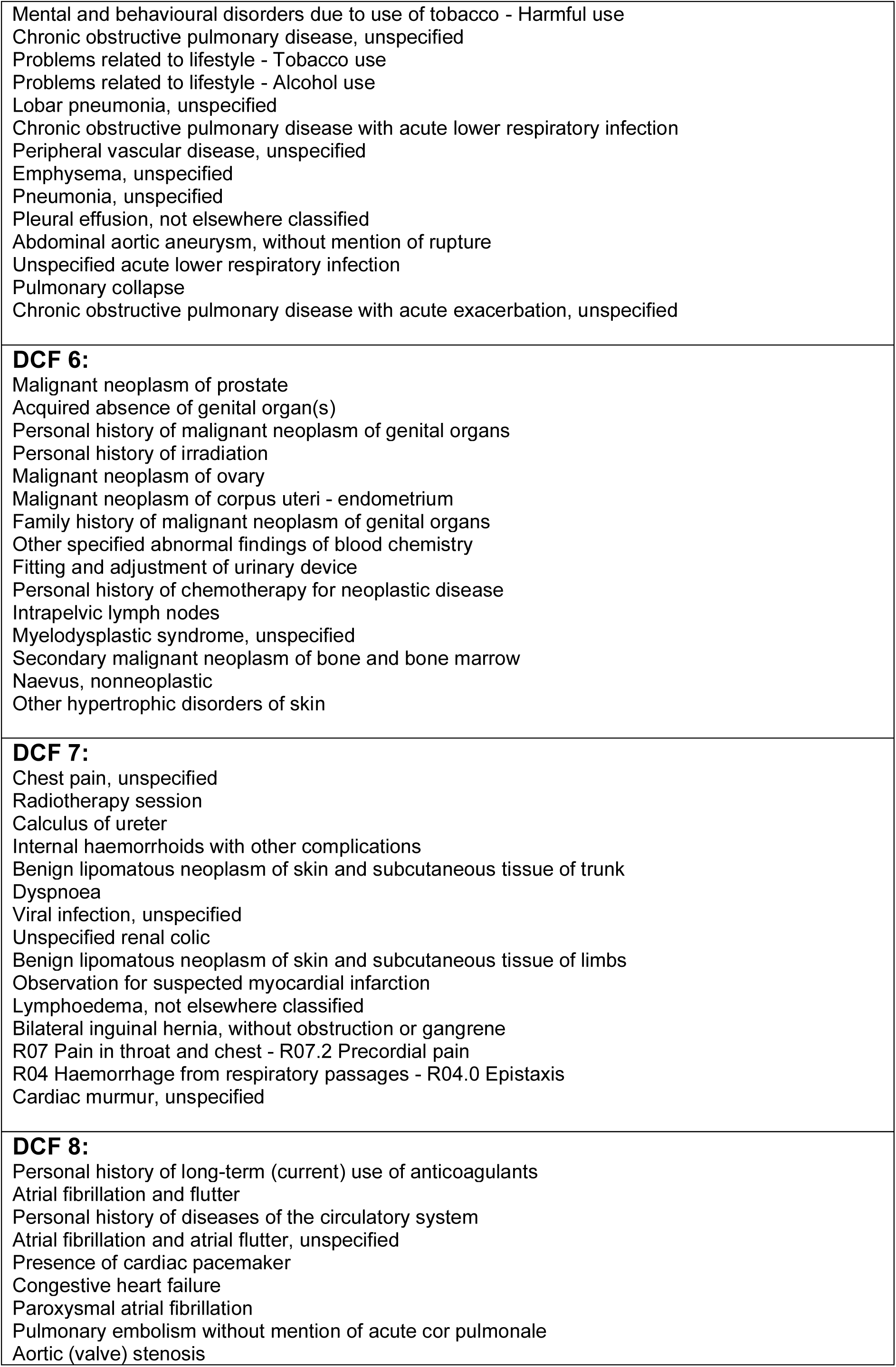
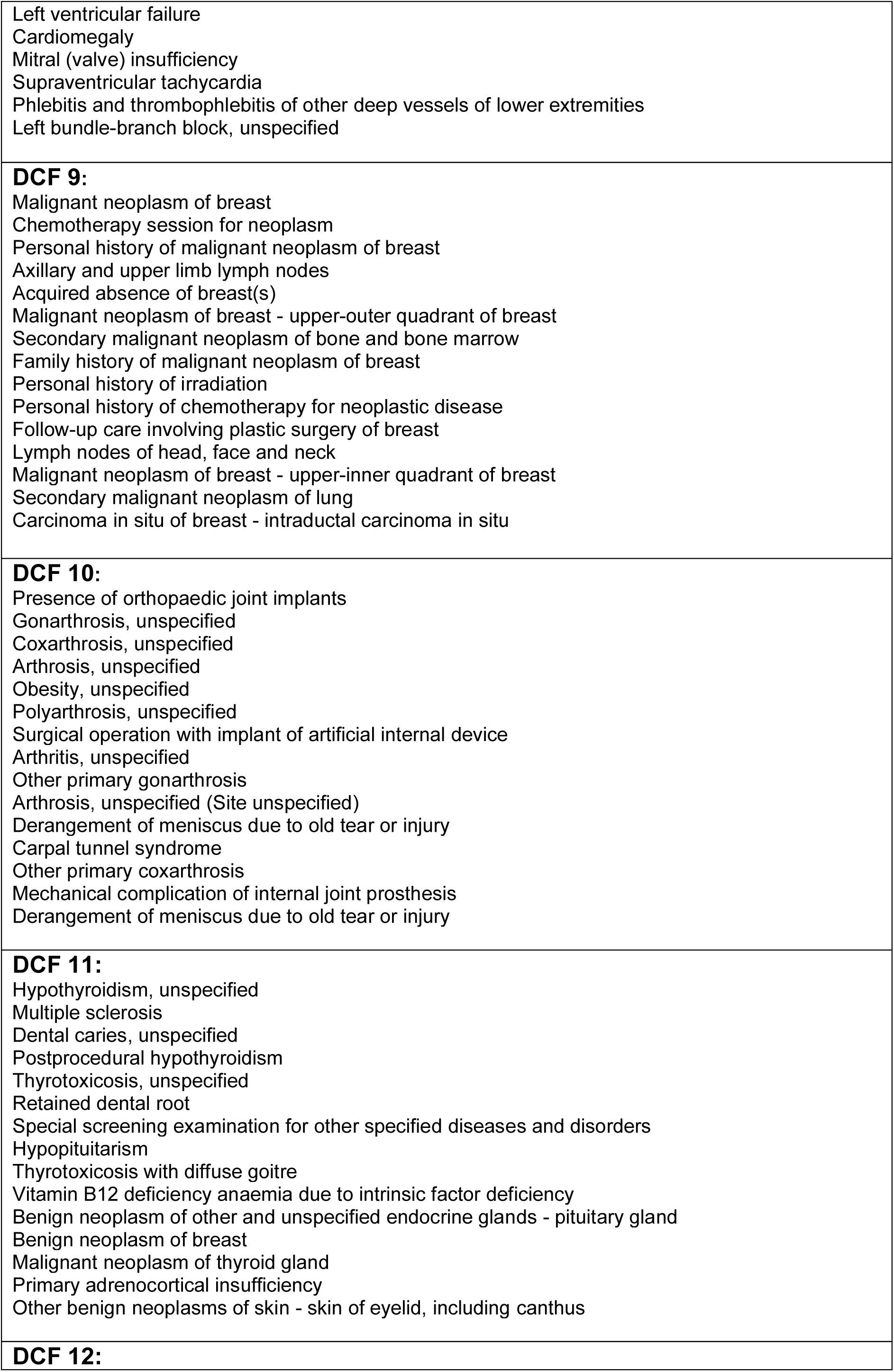
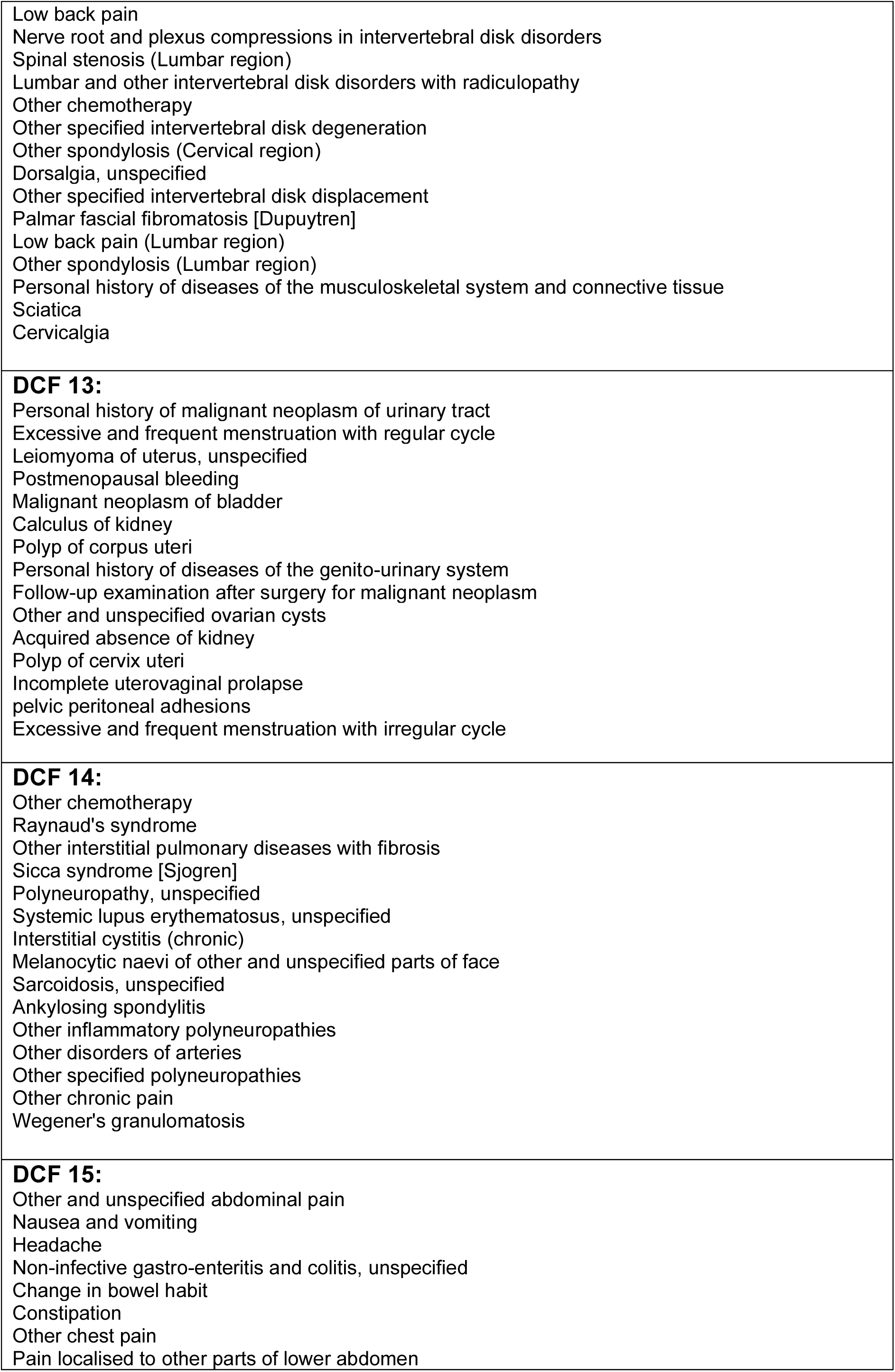
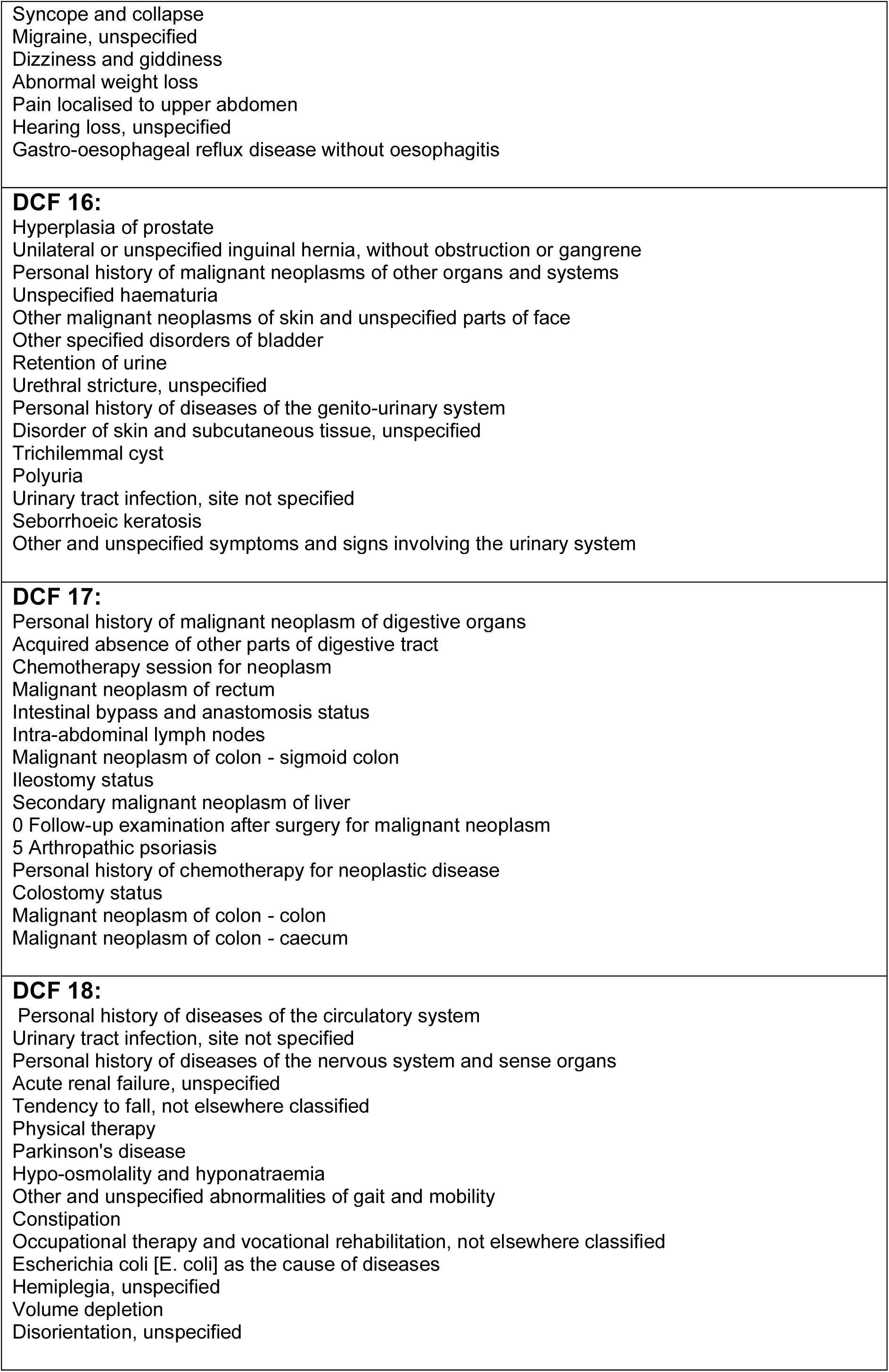
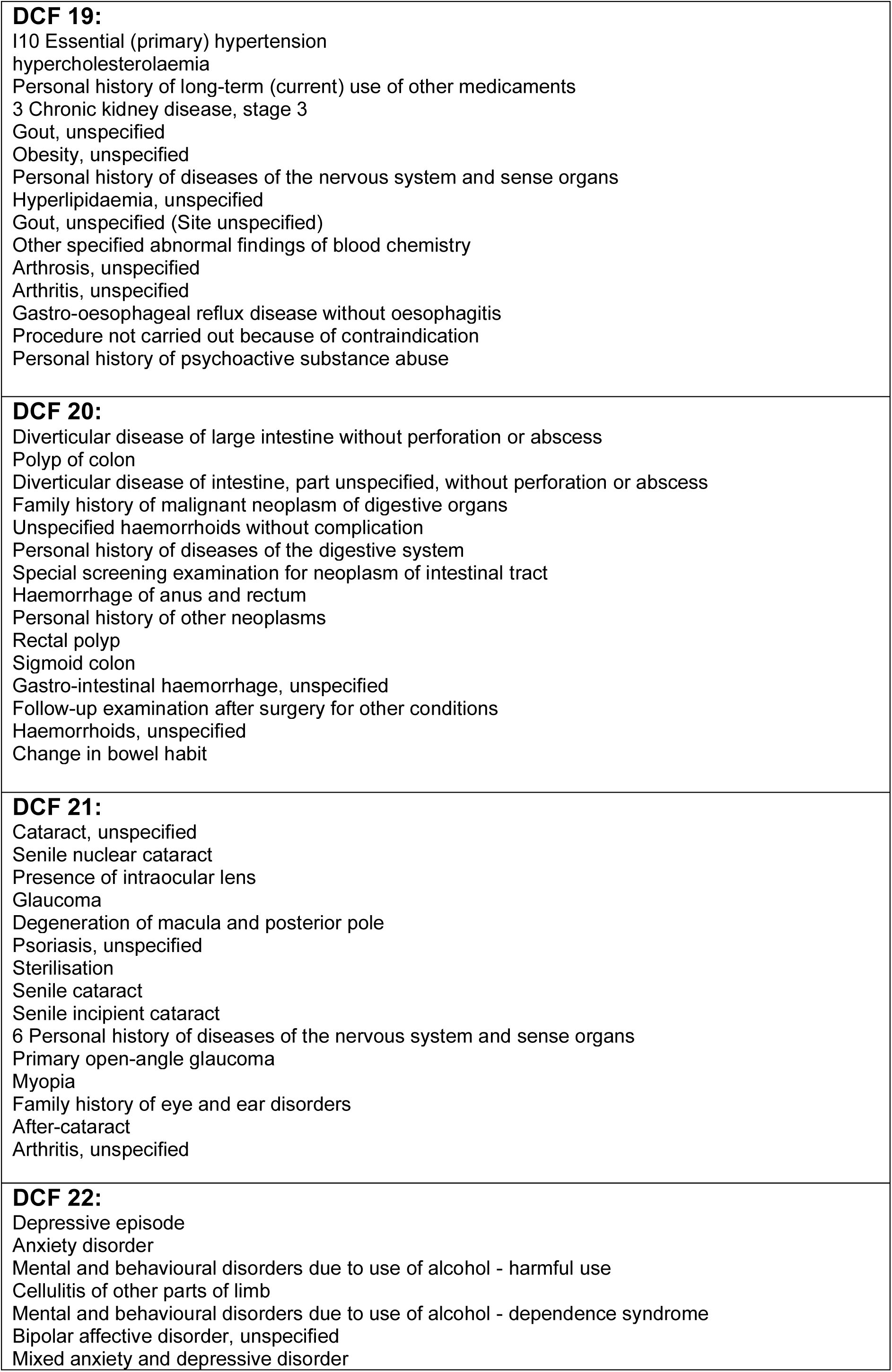
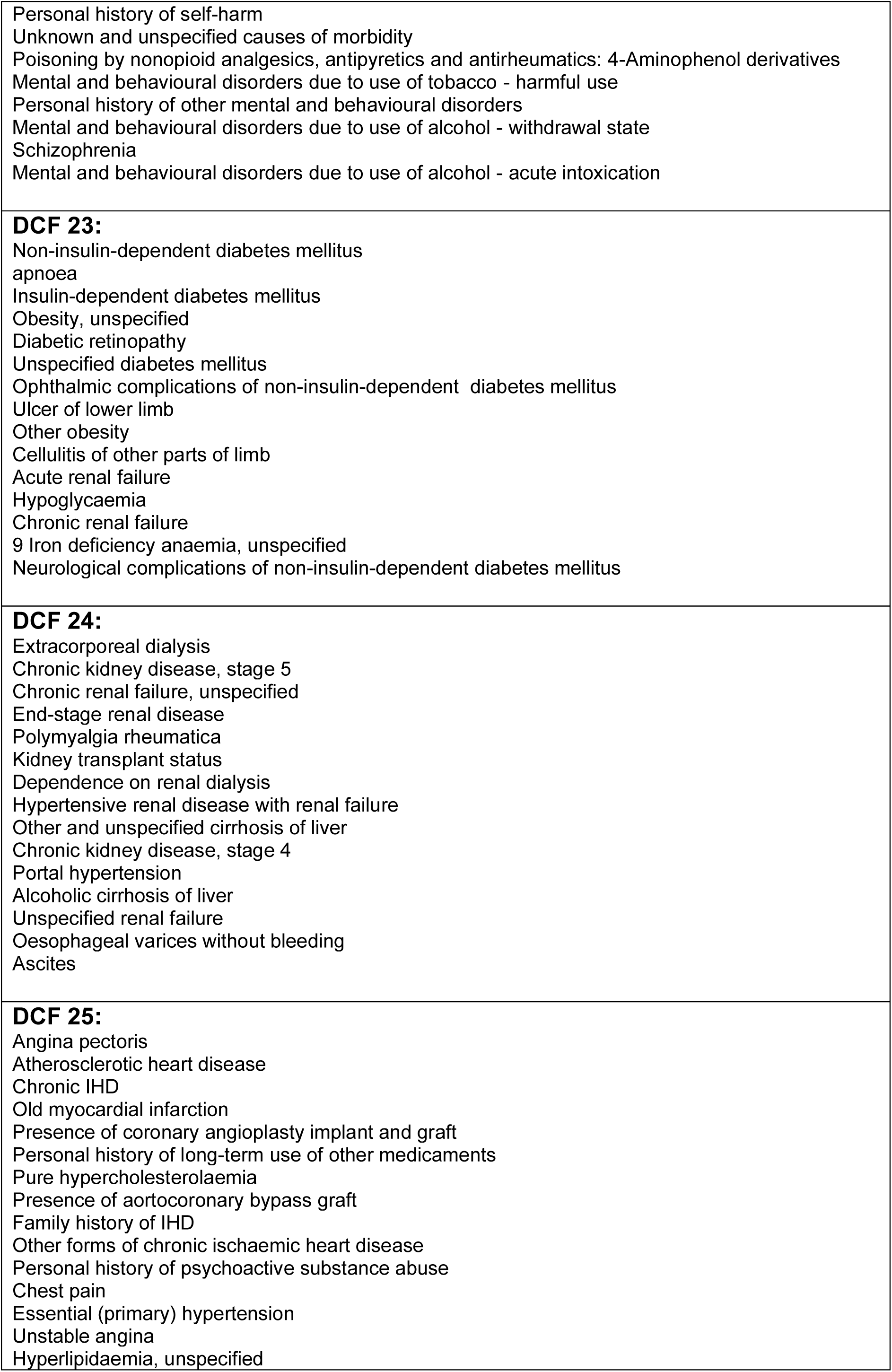
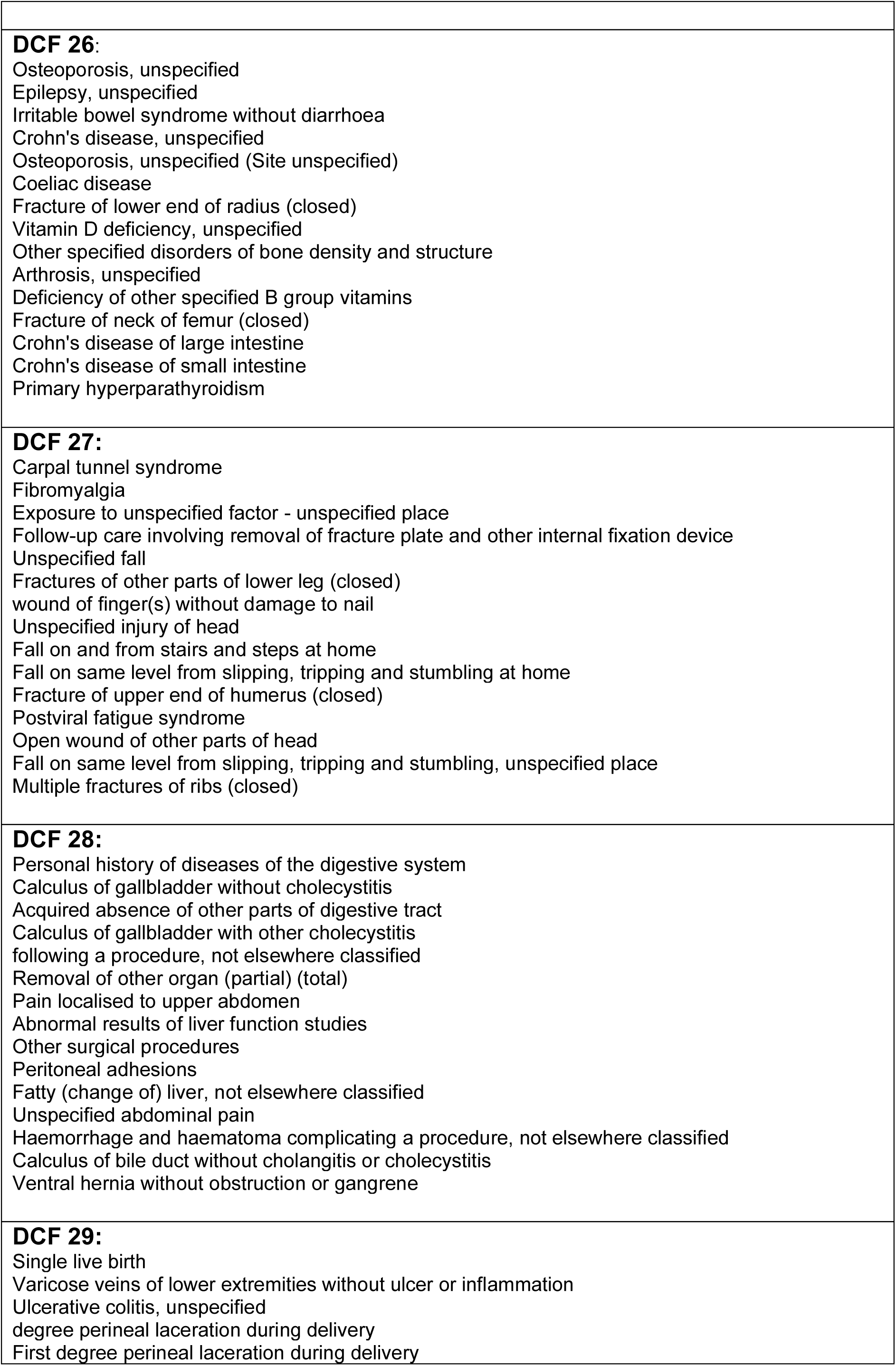
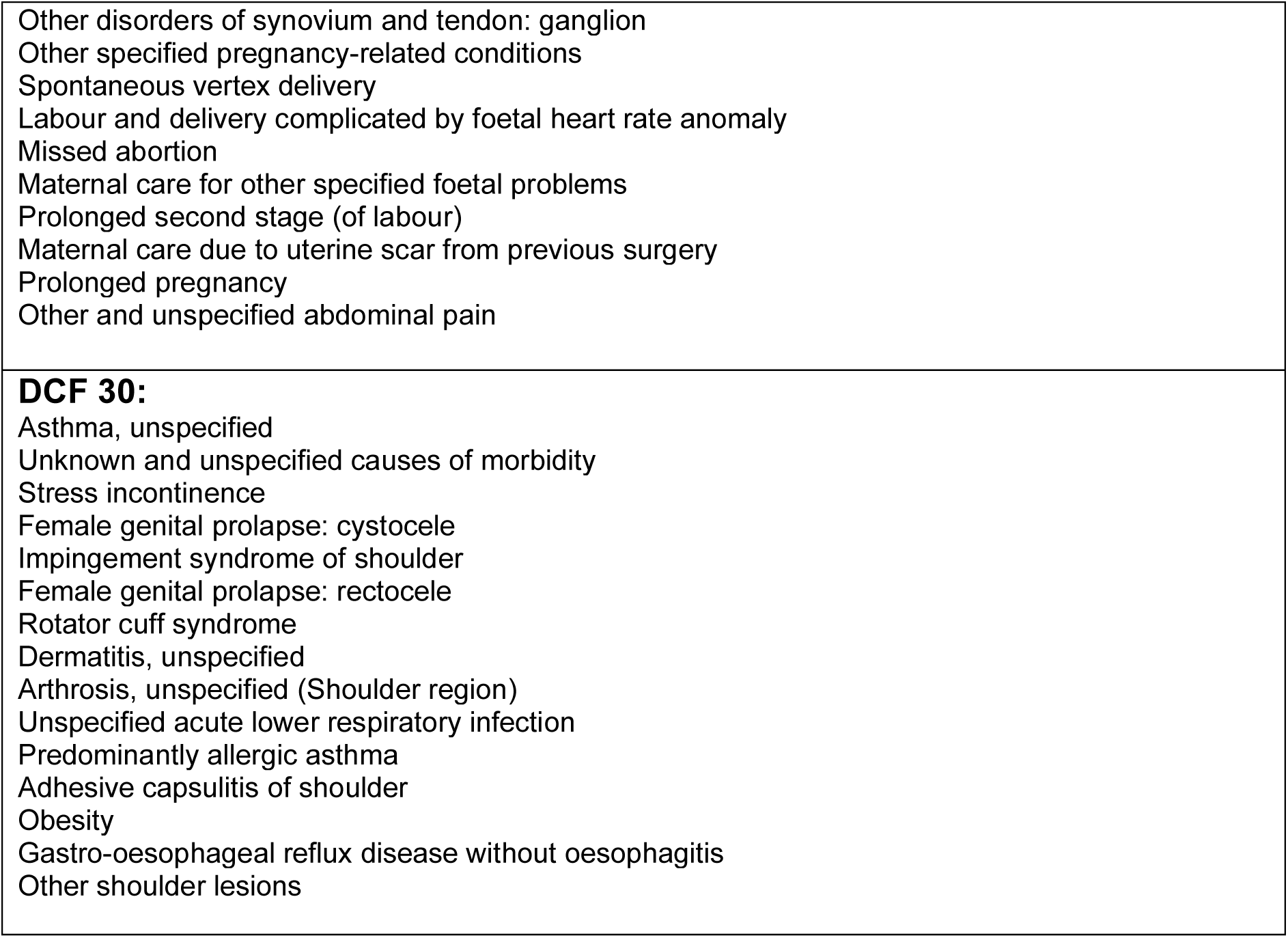
Top 15 disease codes (in rank order) from all 30 topics of the 30-topic LDA model used in the DCF Model

